# Rare variants in long non-coding RNAs are associated with blood lipid levels in the TOPMed Whole Genome Sequencing Study

**DOI:** 10.1101/2023.06.28.23291966

**Authors:** Yuxuan Wang, Margaret Sunitha Selvaraj, Xihao Li, Zilin Li, Jacob A. Holdcraft, Donna K. Arnett, Joshua C. Bis, John Blangero, Eric Boerwinkle, Donald W. Bowden, Brian E. Cade, Jenna C. Carlson, April P. Carson, Yii-Der Ida Chen, Joanne E. Curran, Paul S. de Vries, Susan K. Dutcher, Patrick T. Ellinor, James S. Floyd, Myriam Fornage, Barry I. Freedman, Stacey Gabriel, Soren Germer, Richard A. Gibbs, Xiuqing Guo, Jiang He, Nancy Heard-Costa, Bertha Hildalgo, Lifang Hou, Marguerite R. Irvin, Roby Joehanes, Robert C. Kaplan, Sharon LR. Kardia, Tanika N. Kelly, Ryan Kim, Charles Kooperberg, Brian G. Kral, Daniel Levy, Changwei Li, Chunyu Liu, Don Lloyd-Jone, Ruth JF. Loos, Michael C. Mahaney, Lisa W. Martin, Rasika A. Mathias, Ryan L. Minster, Braxton D. Mitchell, May E. Montasser, Alanna C. Morrison, Joanne M. Murabito, Take Naseri, Jeffrey R. O’Connell, Nicholette D. Palmer, Michael H. Preuss, Bruce M. Psaty, Laura M. Raffield, Dabeeru C. Rao, Susan Redline, Alexander P. Reiner, Stephen S. Rich, Muagututi’a Sefuiva Ruepena, Wayne H-H. Sheu, Jennifer A. Smith, Albert Smith, Hemant K. Tiwari, Michael Y. Tsai, Karine A. Viaud-Martinez, Zhe Wang, Lisa R. Yanek, Wei Zhao, NHLBI Trans-Omics for Precision Medicine (TOPMed) Consortium, Jerome I. Rotter, Xihong Lin, Pradeep Natarajan, Gina M. Peloso

**Affiliations:** Department of Biostatistics, Boston University School of Public Health, Boston, MA, USA; Cardiovascular Research Center and Center for Genomic Medicine, Massachusetts General Hospital, Boston, MA, USA; Program in Medical and Population Genetics, Broad Institute of Harvard and MIT, Cambridge, MA, USA; Department of Medicine, Harvard Medical School, Boston, MA, USA; Department of Biostatistics, Harvard T.H. Chan School of Public Health, Boston, MA, USA; Department of Biostatistics and Health Data Science, Indiana University School of Medicine, Indianapolis, IN, USA; Center for Computational Biology & Bioinformatics, Indiana University School of Medicine, Indianapolis, IN, USA; Provost Office, University of South Carolina, Columbia, SC, USA; Department of Epidemiology and Biostatistics, University of South Carolina Arnold School of Public Health, Columbia, SC, USA; Cardiovascular Health Research Unit, Department of Medicine, University of Washington, Seattle, WA, USA; Department of Human Genetics and South Texas Diabetes and Obesity Institute, University of Texas Rio Grande Valley School of Medicine, Brownsville, TX, USA; Human Genetics Center, Department of Epidemiology, Human Genetics, and Environmental Sciences, School of Public Health, The University of Texas Health Science Center at Houston, Houston, TX, USA; Department of Biochemistry, Wake Forest University School of Medicine, Winston-Salem, NC, USA; Department of Medicine, Brigham and Women’s Hospital, Boston, MA, USA; Division of Sleep Medicine, Harvard Medical School, Boston, MA, USA; Department of Human Genetics, School of Public Health, University of Pittsburgh, Pittsburgh, PA, USA; Department of Biostatistics, School of Public Health, University of Pittsburgh, Pittsburgh, PA, USA; Department of Medicine, University of Mississippi Medical Center, Jackson, MS, USA; The Institute for Translational Genomics and Population Sciences, Department of Pediatrics, The Lundquist Institute for Biomedical Innovation at Harbor-UCLA Medical Center, Torrance, CA, USA; The McDonnell Genome Institute, Washington University School of Medicine, St. Louis, MO, USA; Cardiovascular Research Center, Massachusetts General Hospital, Boston, MA, USA; Cardiovascular Disease Initiative, The Broad Institute of MIT and Harvard, Cambridge, MA, USA; Department of Epidemiology, University of Washington, Seattle, WA, USA; Center for Human Genetics, University of Texas Health at Houston, Houston, TX, USA; Department of Internal Medicine, Nephrology, Wake Forest University School of Medicine, Winston-Salem, NC, USA; Broad Institute of Harvard and MIT, Cambridge, MA, USA; New York Genome Center, New York, NY, USA; Baylor College of Medicine Human Genome Sequencing Center, Houston, TX, USA; Department of Epidemiology, Tulane University School of Public Health and Tropical Medicine, New Orleans, LA, USA; Tulane University Translational Science Institute, New Orleans, LA, USA; Framingham Heart Study, Framingham, MA, USA; Department of Neurology, Boston University Chobanian & Avedisian School of Medicine, Boston, MA, USA; Department of Epidemiology, University of Alabama at Birmingham School of Public Health, Birmingham, AL, USA; Department of Preventive Medicine, Northwestern University, Chicago, IL, USA; Population Sciences Branch, Division of Intramural Research, National Heart, Lung, and Blood Institute, National Institutes of Health, Bethesda, MD, USA; Department of Epidemiology and Population Health, Albert Einstein College of Medicine, Bronx, NY, USA; Division of Public Health Sciences, Fred Hutchinson Cancer Center, Seattle, WA, USA; Department of Epidemiology, University of Michigan, Ann Arbor, MI, USA; Department of Medicine, Division of Nephrology, University of Illinois Chicago, Chicago, IL, USA; Psomagen, Inc. (formerly Macrogen USA), Rockville, MD, USA; GeneSTAR Research Program, Department of Medicine, Johns Hopkins University School of Medicine, Baltimore, MD, USA; The Charles Bronfman Institute for Personalized Medicine, Icahn School of Medicine at Mount Sinai, New York, NY, USA; NNF Center for Basic Metabolic Research, University of Copenhagen, Cophenhagen, Denmark; George Washington University School of Medicine and Health Sciences, Washington, DC, USA; Department of Human Genetics and Department of Biostatistics, University of Pittsburgh, Pittsburgh, PA, USA; Department of Medicine, University of Maryland School of Medicine, Baltimore, MD, USA; Department of Medicine, Boston Medical Center, Boston University Chobanian and Avedisian School of Medicine, Boston, MA, USA; Ministry of Health, Apia, Samoa; Department of Health Systems and Population Health, University of Washington, Seattle, WA, USA; Department of Genetics, University of North Carolina at Chapel Hill, Chapel Hill, NC, USA; Division of Biostatistics, Washington University School of Medicine, St. Louis, MO, USA; Department of Medicine, Brigham and Women’s Hospital, Harvard Medical School, Boston, MA, USA; Center for Public Health Genomics, University of Virginia, Charlottesville, VA, USA; Lutia i Puava ae Mapu i Fagalele, Apia, Samoa; National Health Research Institute (NHRI), Taiwan; Department of Biostatistics, University of Michigan, Ann Arbor, MI, USA; Department of Biostatistics, University of Alabama, Birmingham, AL, USA; Department of Laboratory Medicine and Pathology, University of Minnesota, Minneapolis, MN, USA; Illumina Laboratory Services, Illumina Inc., San Diego, CA, USA; Department of Statistics, Harvard University, Cambridge, MA, USA

## Abstract

Long non-coding RNAs (lncRNAs) are known to perform important regulatory functions. Large-scale whole genome sequencing (WGS) studies and new statistical methods for variant set tests now provide an opportunity to assess the associations between rare variants in lncRNA genes and complex traits across the genome. In this study, we used high-coverage WGS from 66,329 participants of diverse ancestries with blood lipid levels (LDL-C, HDL-C, TC, and TG) in the National Heart, Lung, and Blood Institute (NHLBI) Trans-Omics for Precision Medicine (TOPMed) program to investigate the role of lncRNAs in lipid variability. We aggregated rare variants for 165,375 lncRNA genes based on their genomic locations and conducted rare variant aggregate association tests using the STAAR (variant-Set Test for Association using Annotation infoRmation) framework. We performed STAAR conditional analysis adjusting for common variants in known lipid GWAS loci and rare coding variants in nearby protein coding genes. Our analyses revealed 83 rare lncRNA variant sets significantly associated with blood lipid levels, all of which were located in known lipid GWAS loci (in a ±500 kb window of a Global Lipids Genetics Consortium index variant). Notably, 61 out of 83 signals (73%) were conditionally independent of common regulatory variations and rare protein coding variations at the same loci. We replicated 34 out of 61 (56%) conditionally independent associations using the independent UK Biobank WGS data. Our results expand the genetic architecture of blood lipids to rare variants in lncRNA, implicating new therapeutic opportunities.

## Introduction

Blood lipid levels, including low-density lipoprotein cholesterol (LDL-C), total cholesterol (TC), triglyceride (TG), high-density lipoprotein cholesterol (HDL-C), are quantitative clinically important traits with well-described monogenic and polygenic bases^1–19^. Abnormal blood lipid levels contribute to risk of coronary heart disease (CHD) and, in clinical practice, several treatments, including statins, PCSK9 and ANGPTL3 inhibitors^20–22^, are available to reduce the risk of developing CHD. Each of these therapeutics has supporting evidence of their efficacy from human genetic analysis of blood lipid levels^21–23^.

Long non-coding RNAs (lncRNAs) are broadly defined as transcripts greater than 200 nucleotides in length that biochemically resemble mRNAs but do not code for proteins^24^. lncRNAs are known to perform important regulatory functions in lipid metabolism^25–27^. Rare variants (RVs) in lncRNAs have not been systematically explored for their impact on blood lipid levels as they are not comprehensively genotyped or imputed on non-WGS platforms. In addition, there are difficulties in defining testing units and selecting qualifying variants^28^. Rapidly growing knowledge about the regulatory elements of the non-coding genome^29–33^, large-scale WGS studies ^34–36^, and new statistical methods ^37–39^ for variant set tests provide the possibility to assess the associations between plasma lipid traits and the genome-wide impact of lncRNAs.

We examined the associations of rare variants in lncRNA genes from high-coverage WGS of 66,329 participants from diverse ancestry who have blood lipid traits (LDL-C, HDL-C, TC and TG) in the National Heart, Lung, and Blood Institute (NHLBI) Trans-omics for Precision Medicine (TOPMed) program freeze 8 data^34^. We show that the rare noncoding variants in lncRNA genes located near known Mendelian dyslipidemia genes contribute to phenotypic variation in lipid levels among unselected individuals from population-based cohorts biobanks independently of common variants associated with blood lipid levels.

## Results

### Overview

We performed a comprehensive evaluation of the association between quantitative blood lipid traits and rare variants in lncRNA genes across the genome (Figure 1). We systematically curated more than 165k lncRNA genes from the union of four human genome lncRNA annotations, including GENCODE ^29,30^, FANTOM5 CAT^31^, NONCODE^32^ and lncRNAKB^33^. We utilized the TOPMed Freeze 8 dataset of 66,329 participants from 21 studies with WGS and measured blood lipid levels and performed the rare variant (MAF <1%) association tests of curated lncRNA genes with four blood lipid phenotypes: LDL-C, HDL-C, TC, and TG. We further conducted the conditional analysis adjusting for known genome-wide association study (GWAS) variants from the Global Lipids Genetics Consortium (GLGC)^18^. Associations between lncRNA genes and lipids that were conditionally independent from the GWAS variants (conditional *P* value < 6.0e-04) were then tested using STAAR procedure for conditional analysis adjusting for rare nonsynonymous variants (MAF < 1%) within the closest protein coding gene and the nearby known lipid monogenic genes in the region. We performed replication in ∼140□K genomes from UK Biobank^40^. We intersected our results with the gene expression signatures of lipid traits in 1,505 participants from the Framingham Heart Study (FHS)^41^ with RNA-seq data and blood lipid levels and observed evidence that the lncRNA RVs may both influence their gene expression levels and impact lipid traits.

**Figure 1.**
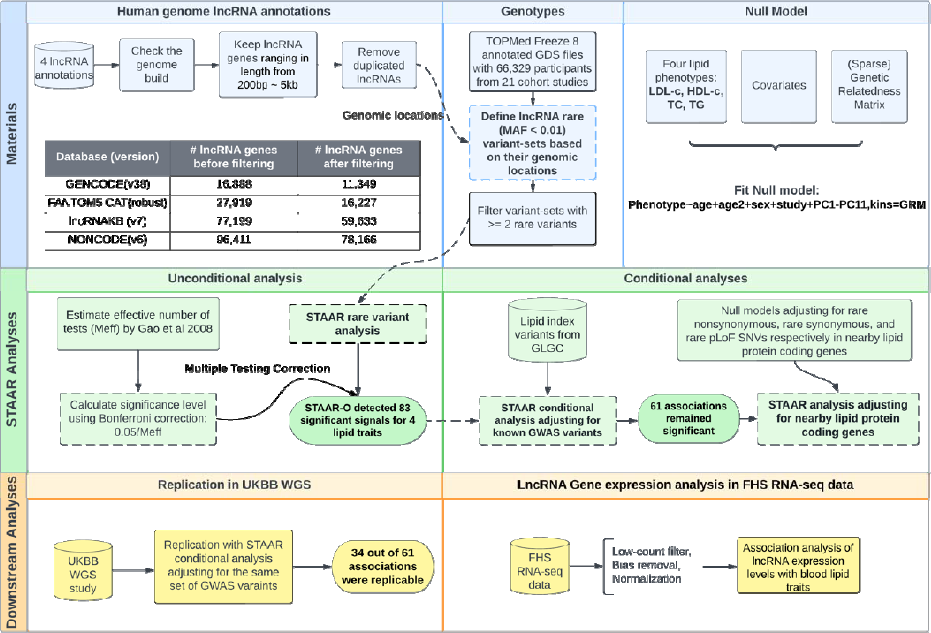
A schematic illustration of the study.

### Characteristics of TOPMed participants

We included 66,329 diverse participants from 21 cohort studies in the NHLBI TOPMed consortium with blood lipid levels. The discovery cohorts consisted of 29,502 (44.5%) self-reported White, 16,983 (25.6%) self-reported Black, 13,943 (21.0%) self-reported Hispanic, 4,719 (7.1%) self-reported Asian, and 1,182 (1.8%) self-reported Samoan participants (**Supplementary Table 1, Supplementary Text**). Among the 66,329 participants, 41,182 (62%) were female. The mean age of the 66,329 participants was 53 years (SD = 15). The mean ages at lipid measurement varied across 21 cohorts from 25 years (SD = 3.56) for the Coronary Artery Risk Development in Young Adults (CARDIA) to 73 years (SD = 5.38) for the Cardiovascular Health Study (CHS). We observed that the Amish cohort had a higher concentration of LDL-C (140 [SD = 43] mg/dL) and HDL-C (56 [SD = 16] mg/dL) as well as lower TG (median 63 [IQR = 50] mg/dL) consistent with the known founder mutations in *APOB* and *APOC3*^35^.

### Identification of rare lncRNA variants associated with blood lipid traits

We defined lncRNA testing units using the available genomic positions in four genome annotation projects described in the **Methods**. There were 11,349 lncRNA genes obtained from GENCODE^29,30^, 16,227 from FANTOM5 CAT^31^, 78,166 from NONCODE^32^ and 59,633 from lncRNAKB^33^. In total, we tested 165,375 lncRNA genes, among which, the average number of rare variants in each lncRNA was 483 (SD = 572). The minimum and the maximum number of rare variants among the lncRNAs being tested are 2 and 2947, respectively.

Our aggregation of lncRNAs across four lncRNA resources led to an overlap in the lncRNA units, leading to non-independent tests of association of the lncRNAs with blood lipid levels. We estimated the effective number of tests (*M_eff_*) using a principal component analysis (PCA) based approach^42^ since the traditional Bonferroni correction would be too conservative and reduce power to detect association with blood lipid levels^28^. *M_eff_* was estimated as 111,550, providing a significance threshold of a = 0.05/111,550 = 4.5 X10^−7^.

We applied STAAR (variant-Set Test for Association using Annotation infoRmation) framework^37,38^ to identify the lncRNA rare variant (RV) sets that associated with quantitative lipid traits (LDL-C, HDL-C, TC and TG) using TOPMed WGS data. STAAR-O identified 83 genome-wide significant associations (28 with LDL-C, 20 with TC, 19 with HDL-C, and 16 with TG) (Table 1, **Supplementary Table 2)**. Among the 83 genome-wide significant associations, there are 54 unique lncRNAs. We observed that all the significant associations in the unconditional analysis were in the known lipid GWAS loci (defined as a ±500 kb window beyond a Global Lipids Genetics Consortium index variant)^18^. We performed a sensitivity analysis aggregating only exonic and splicing variants in lncRNA genes and observed consistent results to our primary analysis results (**Supplementary Figure 1**).

**Table 1.** Summary of significant lncRNA associations for unconditional analysis, conditional analyses, and replication.

| Method | LDL-C | TC | HDL-C | TG | Total No. |
| --- | --- | --- | --- | --- | --- |
| **STAAR Unconditional analysis\*** | 28 | 20 | 19 | 16 | 83 |
| **Conditioning on known lipid-associated variants \*\*** | 20 | 14 | 15 | 12 | 61 |
| **Conditioning on rare nonsynonymous variants within the closest gene and nearby lipid monogenic genes \*\*\*** | 18 | 13 | 15 | 12 | 58 |
| **Conditioning on rare synonymous variants within the closest gene and nearby lipid monogenic genes \*\*\*** | 20 | 14 | 15 | 12 | 61 |
| **Conditioning on rare pLoF variants within the closest gene and nearby lipid monogenic genes \*\*\*** | 20 | 14 | 15 | 12 | 61 |
| **Replication in UKBB WGS \*\*\*** | 13 | 7 | 8 | 6 | 34 |

\* Bonferroni correction level of $0.05/111,550 = 4.5e-07$
\*\*Bonferroni correction level of $0.05/83 = 6.0e-04$
\*\*\*Bonferroni correction level of $0.05/61 = 8.2e-04$

### Conditional analyses of trait-associated lncRNAs adjusting for known GWAS variants and nonsynonymous variants within the nearby lipid monogenic genes

After conditioning on known lipid-associated variants in a ±500 kb window beyond a variant set^18^, 61 out of 83 associations (73%) remained significant (20 with LDL-C, 14 with TC, 15 with HDL-C, and 12 with TG) at the Bonferroni corrected level of 0.05/83 = 6.0 × 10^−4^, indicating that the associations between the lncRNA genes and lipid levels are distinct from the known GWAS variants. The most significant association for LDL-C and TC was the lncRNA NONHSAG026007.2 (chr19:44,892,420-44,903,056) near the *APOE-APOC1* region. NONHSAG026007.2 remained significantly associated with LDL-C (*P* value = 2.44 × 10^−15^) and TC (*P* value =2.17 × 10^−27^) after adjusting for nearby known lipid-associated variants (Figure 2). The most significant associations for HDL-C and TG were NONHSAG063125.1 (chr11:116,790,241-116,805,983) and NONHSAG09700.3 (chr11: 116,773,068-116,779,841), respectively, both near *APOA5-APOC3-APOA1* region. NONHSAG063125.1 remained similarly associated after conditioning on known lipid GWAS variants, while NONHSAG09700.3 became even more significant (Figure 2). We then conditioned the GWAS-distinct associations on the rare nonsynonymous variants within the closest protein coding gene and nearby lipid monogenic genes and observed that most (94.9%) of the lncRNA associations with lipid levels remained significant (Table 1; **Supplementary Figure 2**). Additionally, when conditioned on the rare synonymous variants or rare pLoF variants within the closest protein coding gene and nearby lipid monogenic genes, the number of associations remained as same as those GWAS-distinct associations (Table 1; **Supplementary Figure 3**).

**Figure 2.**
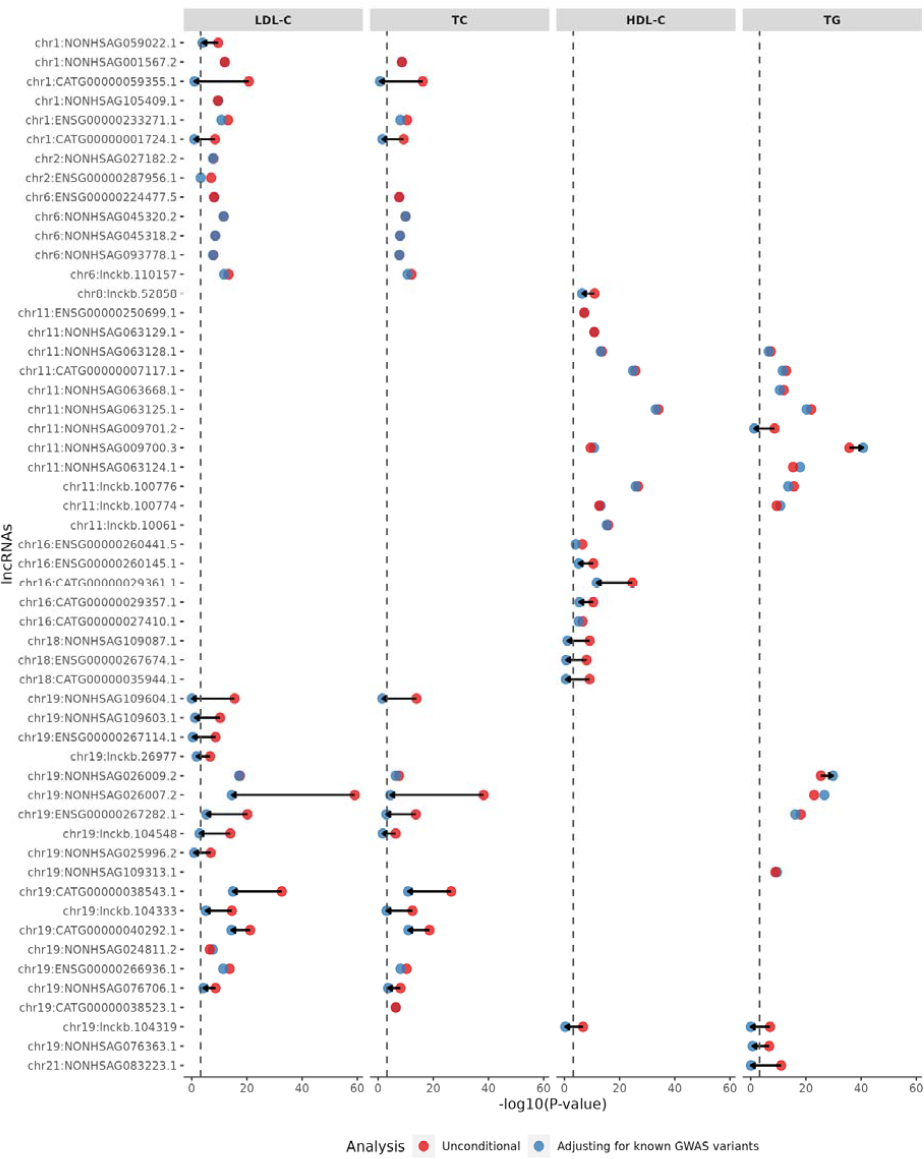
Significantly associated lncRNAs with four blood lipid traits (STAAR-O *P* value < 4.5e-07) The lncRNA genes are ordered by chromosome, followed by genomic positions. Dots in red and blue represent the −log_10_(STAAR-O *P* value) of the STAAR unconditional and conditional analysis adjusting for known lipid-associated GWAS variants, respectively. The black dashed line is the Bonferroni correction level of 0.05/83 = 6.0e-04. Arrows indicate at least 10^4^ fold change of STAAR-O *P* values comparing the unconditional analysis and conditional analysis adjusting for known lipid-associated GWAS variants.

### Replication of significant lncRNA-blood lipid trait associations

Replication of 61 lncRNAs associated with blood lipid levels was evaluated in 139,849 UK Biobank individuals with WGS and blood lipid levels (**Supplementary Table 3)**. We replicated 34 out of 61 (56%) lncRNA associations with blood lipid levels at a Bonferroni-corrected threshold of 0.05/61 = 8.2e-04 (**Supplementary Table 2**). The most significant associations in the UK Biobank replication were NONHSAG025996.2 (chr19: 44,694,720-44,696,054) near *APOE-APOC1* region for LDL-C, NONHSAG109604.1 near *APOE-APOC1* region for TC, NONHSAG009700.3 near *APOA5-APOC3-APOA1* region for both HDL-C and TG (**Supplementary Table 2**), which were consistent with the results from TOPMed.

### lncRNA gene expression analysis in FHS RNA-seq data

We overlapped the significant lipid-associated lncRNA genes with the lncRNA genes available in the Framingham Heart Study (FHS) RNA-seq data generated by TOPMed^43^. Since the gene-level expression data in FHS is annotated by GENCODE v30, we limited the lncRNA genes to those presented in GENCODE. Among the 54 unique lncRNA genes that are significantly associated with either one of the lipid traits using TOPMed WGS data, 10 lncRNA genes are annotated by GENCODE, and 8 out of 10 can be found in the FHS data. We performed association analyses of expression levels of those 8 significant lipid-associated lncRNA genes with blood lipid levels (LDL-C, TC, HDL-C, TG) (**Supplementary Text, Supplementary Table 4**). In total, we tested 12 associations of lncRNA gene expression with blood lipid level (**Supplementary Table 4**). The small proportion of overlapping was partially due to lncRNA genes’ generally lower expression. The lowly expressed genes were filtered out when processing the gene expression data.

Four associations achieved Bonferroni-adjusted significance, including the gene expression level of ENSG00000267282.1 (chr19:44,881,088-44,890,922) associated with LDL-C, TC, and TG, and the gene expression level of ENSG00000266936.1 (chr19:11,010,917-11,016,011) associated with TC. ENSG00000267282.1 is an antisense of *NECTIN2* (also known as *PVRL2*) (Figure 3). The nectin cell adhesion molecule 2 (*NECTIN2*) protein is a cell adhesion molecule involved in lipid metabolism^44^. Additionally, ENSG00000267282.1 was one of the lncRNA associations that we replicated in the independent UK Biobank (**Supplementary Table 2**). We also queried whether the RVs in this lipid-associated lncRNA led to an alteration of the corresponding lncRNA levels in the blood. However, due to the small number of overlapping individuals between FHS RNA-seq data and TOPMed WGS data (N = 512), the number of RVs tested in ENSG00000267282.1 for the association of its gene expression level was only 59. Compared with the original analysis using all 66,329 individuals for the association with lipid levels, the number of RVs tested in ENSG00000267282.1 is 1417. As a result, the association of the RVs in the ENSG00000267282.1 with ENSG00000267282.1 gene expression levels in blood was not significant (STAAR-O *P* value = 0.68).

**Figure 3.**
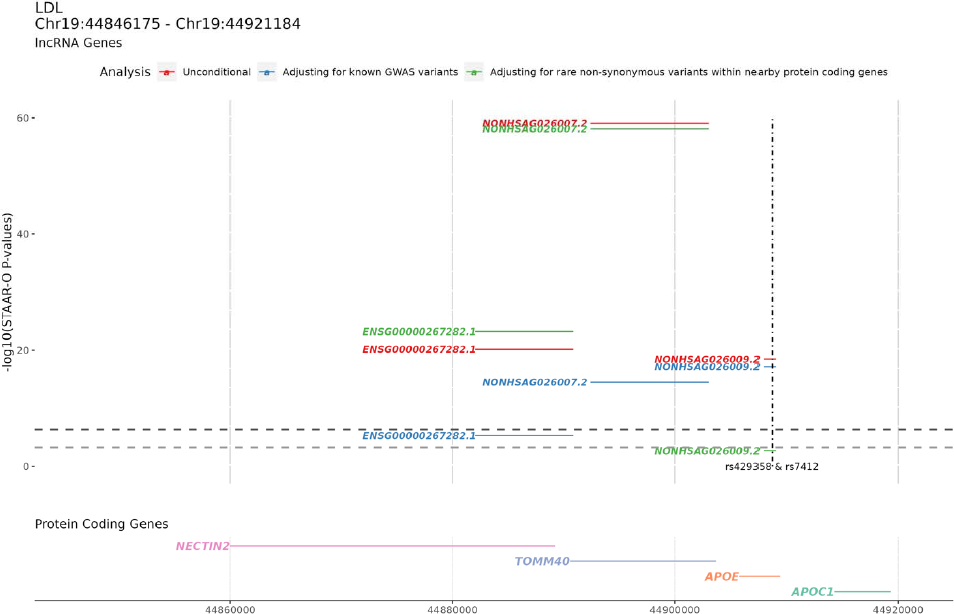
lncRNAs in the APOE region associated with LDL-C. Upper panel shows the - log_10_(STAAR-O *P* value) of the STAAR unconditional analysis, STAAR conditional analysis adjusting on known lipid GWAS variants, and STAAR conditional analysis adjusting for rare non-synonymous variants within the closest protein-coding gene and nearby lipid monogenic genes. The bottom panel is the nearby protein coding genes with the genomic coordinates. The vertical dashed line is the position of the known GWAS variants that were conditioned on. The black horizontal dashed line is the Bonferroni correction level of 0.05/111,550 = 4.5e-07, and the gray horizontal dashed line is the Bonferroni correction level of 0.05/83 = 6.0e-04.

## Discussion

In this study, we conducted genome-wide rare-variant associations of 165K lncRNAs in ancestrally diverse TOPMed participants (*N* = 66,329) with measured blood lipid levels. Using rare-variant association tests, we observed 83 rare lncRNAs significantly associated with blood lipid levels, and of these, 61 (73%) were conditionally distinct from common regulatory variation and rare protein coding variation at the same loci. Notably, most of these association signals were replicated in an independent WGS dataset, UK Biobank. We also highlighted one trait-associated lncRNA, ENSG00000267282.1(chr19:44,881,088-44,890,922), whose gene expression level was also shown to be associated with lipid levels using RNA-seq data from the FHS. Together, this systematic assessment of rare lncRNA variants suggests an additional genomic element in known lipid gene regions that is distinct from the known lipid genes.

Genetic variation for blood lipids levels has been observed across the allelic spectrum with common, rare coding, and rare non-coding variants being associated with blood lipids levels^36^. Blood lipids have been associated with non-coding regulatory variants and coding variation in genes, and now also associated with lncRNAs. We show that all the trait-associated lncRNAs are in genomic regions previously associated with blood lipid traits, leading to the plausibility of these results. About 75% of the associations are conditionally distinct from common regulatory variation and rare protein coding variation at the same loci previously identified through GWAS and whole exome sequencing studies. This indicates that the regulatory variants through lncRNAs additionally contribute to the variation of blood lipid levels.

Despite numerous reports indicating the potential regulatory role of long non-coding RNAs (lncRNAs), only a small proportion of them have substantial evidence to support such claims^25,26,45^. The fraction of lncRNAs that are functional remains unknown. Through a comprehensive study of over 165,000 lncRNAs, we found that the majority of lncRNAs are not associated with a lipid trait, which aligns with the argument made previously that only a few human lncRNAs contribute centrally to human physiology^45^. However, there are still some lncRNAs that harbor variants that predispose individuals to phenotypic differences in blood lipid levels. Our results suggest that investigators should first prioritize individual lncRNAs near the known trait-associated loci for analysis, which is more likely to yield robust experimental observations.

We further investigated one lncRNA, liver-expressed liver X receptor-induced sequence (*LeXis*), which is a mediator of the complex effects of liver X receptor (LXR) signaling on hepatic lipid metabolism to maintain hepatic sterol content and serum cholesterol levels^46,47^. A potential orthologue of *LeXis* in humans, TCONS_00016452 (chr9:104,990,086-104,991,780), is found in a region adjacent to the human *ABCA1* gene. It didn’t stand out as a significant signal for any lipid trait in our study, which might suggest that it was not a functional orthologue of *LeXis*. However, the rapid evolutionary turnover of lncRNAs still hinders the functional identification between species^45,47^.

Several limitations of our study should be noted. First, our RNA-seq analyses were restricted to GENCODE annotation. The small proportion of overlapping RNA-seq data and WGS data limits the ability to test rare lncRNA variants with their gene expression. Second, we did not correct for the number of tested lipid traits however, there is a moderate to high correlation among the blood lipid levels and therefore this would lead to over correction. Third, to assess a causal role of the rare lncRNA variants, we need to further show that they are correlated with lncRNA expression but not correlated with altered expression or function of other genes nearby.

In summary, our results from a large ancestrally diverse participants add further evidence that lncRNA is an additional genomic element in known lipid gene regions that is distinct from the known genes. We comprehensively evaluated 165K lncRNAs for their association with variation in lipid traits and replicated most of the signals in an independent UKB WGS cohort.

## Methods

### Discovery and replication cohorts

#### Discovery cohorts

The discovery cohort included 66,329 participants in the NHLBI Trans-Omics for Precision Medicine (TOPMed) from 21 cohort studies with Freeze 8 whole genome sequencing (WGS) and blood lipid levels available: Old Order Amish (Amish; n□=□1083), Atherosclerosis Risk in Communities study (ARIC; n□=□8016), Mt Sinai BioMe Biobank (BioMe; n□=□9848), Coronary Artery Risk Development in Young Adults (CARDIA; n□=□3,056), Cleveland Family Study (CFS; n□=□579), Cardiovascular Health Study (CHS; n□=□3,456), Diabetes Heart Study (DHS; n□=□365), Framingham Heart Study (FHS; n□=□3992), Genetic Studies of Atherosclerosis Risk (GeneSTAR; n□=□1757), Genetic Epidemiology Network of Arteriopathy (GENOA; n□=□1046), Genetic Epidemiology Network of Salt Sensitivity (GenSalt; n□=□1772), Genetics of Lipid-Lowering Drugs and Diet Network (GOLDN; n□=□926), Hispanic Community Health Study - Study of Latinos (HCHS-SOL; n□=□7714), Hypertension Genetic Epidemiology Network and Genetic Epidemiology Network of Arteriopathy (HyperGEN; n□=□1853), Jackson Heart Study (JHS; n□=□2847), Multi-Ethnic Study of Atherosclerosis (MESA; n□=□5290), Massachusetts General Hospital Atrial Fibrillation Study (MGH_AF; n□=□683), San Antonio Family Study (SAFS; n□=□619), Samoan Adiposity Study (Samoan; n□=□1182), Taiwan Study of Hypertension using Rare Variants (THRV; n□=□1982) and Women’s Health Initiative (WHI; n□=□8263). The discovery cohorts consisted of 29,502 (44.5%) White, 16,983 (25.6%) Black, 13,943 (21.0%) Hispanic, 4719 (7.1%) Asian, and 1182 (1.8%) Samoan. More information for study descriptions can be found in **Supplementary Table 1**.

#### Replication cohorts

We sought to replicate the findings using the UK Biobank WGS data for 139,849 genomes with blood lipid traits^40^. The UK Biobank is a large, population-based prospective cohort of half a million United Kingdom residents aged 40–69 years. The replication cohorts consisted of 116, 335 White, and 23,335 others **(Supplementary Table 3).**

#### Ethical regulations

Participants from each of the studies contributing to the NHLBI TOPMed consortium provided informed consent, and all studies were approved by IRBs in each of the participating institutions.

### TOPMed WGS Freeze 8 data

#### Phenotype data

We included four conventionally measured blood lipids in this study: low-density lipoprotein cholesterol (LDL-C), total cholesterol (TC), triglyceride (TG), high-density lipoprotein cholesterol (HDL-C). Detailed phenotype calculation and harmonization were described elsewhere^36^. Briefly, LDL-C was either directly measured or calculated by the Friedewald equation when triglycerides were <400□mg/dL. We adjusted the total cholesterol by dividing by 0.8 and LDL-C by dividing by 0.7 when statins were present^10,35^. For triglycerides, we additionally performed the natural log transformation for analysis, since triglycerides were skewed. We then fitted a linear regression model for each phenotype to get the residuals after adjusting for age, age2, sex, race/ethnicity, study and the first 11 ancestral PCs (as recommended by the TOPMed DCC). For Amish participants, we additionally adjusted for APOB p.Arg3527Gln in LDL-C and TC, and adjusted for APOC3 p.Arg19Ter in HDL-C and TG^48–50^. The residuals were inverse rank normalized and rescaled by the standard deviation of the original phenotype within each group^36^.

#### Genotype data

Whole genome sequencing data were accessed from the TOPMed Freeze 8 release. DNA samples were sequenced at the >30× target coverage at seven centers (Broad Institute of MIT and Harvard, Northwest Genomics Center, New York Genome Center, Illumina Genomic Services, PSOMAGEN [formerly Macrogen], Baylor College of Medicine Human Genome Sequencing Center, and McDonnell Genome Institute [MGI] at Washington University)^34^. The reads were aligned to human genome build GRCh38 using the BWA-MEM algorithm. The genotype calling was performed using the TOPMed variant calling pipeline (https://github.com/statgen/topmed_variant_calling). The resulting BCF files were converted to SeqArray GDS format and annotated were annotated internally by curating data from multiple database sources using Functional Annotation of Variant–Online Resource (FAVOR (http://favor.genohub.org)^37,39^. The resulting annotated GDS (aGDS) files were used in this study. We computed the genetic relationship matrix (GRM) using R package *PC-relate* and subtracted GRM of those samples with lipid phenotypes using R package *GENESIS*.

### Human reference genome annotations for long non-coding RNA genes

Multiple lncRNA annotations are available. We obtained four long non-coding RNAs (lncRNAs)annotation resources with different qualities and sizes and merged them to improve comprehensiveness. They included GENCODE ^29,30^, FANTOM5 CAT^31^, NONCODE^32^ and lncRNAKB^33^.

#### GENCODE

GENCODE is the default human reference genome annotation for both Ensembl and UCSC genome browsers. It is also widely adopted by many large-scale genomic consortiums including TOPMed. GENCODE gene sets cover lncRNAs, pseudogenes and small RNAs in addition to protein-coding genes. The lncRNA annotation in GENCODE is almost entirely manual, which ensures the quality and consistency of the data. We downloaded the GENCODE v38 (December 2020) human release from https://ftp.ebi.ac.uk/pub/databases/gencode/Gencode_human/release_38/gencode.v38.long_noncoding_RNAs.gtf.gz, and kept 17,944 lncRNAs genes with a stable identifier and the genomic location information.

#### FANTOM CAT

The Functional Annotation of the Mammalian genome (FANTOM) CAGE-associated transcriptome (CAT) meta-assembly combines both published sources and in-house short-read assemblies. It utilized CAGE tags, which mark transcription start sites (TSSs), to identify human lncRNA genes with high-confidence 5’ ends. We acquired the FANTOM CAT (lv3 robust) lncRNAs assembly from https://fantom.gsc.riken.jp/5/suppl/Hon_et_al_2016/data/assembly/lv3_robust/FANTOM_CAT.lv3_robust.only_lncRNA.gtf.gz. Since the FANTOM5 annotations were on genome version hg19 (GRCh37), we lifted over to genome version hg38 (GRCh38) using the UCSC liftOver tool^51^.

#### lncRNAKB

Long non-coding RNA Knowledgebase (lncRNAKB) is an integrated resource for exploring lncRNA biology in the context of tissue-specificity and disease association. A systematic integration of annotations using a cumulative stepwise intersection method from six independent databases resulted in 77,199 human lncRNA. We downloaded the lncRNAKB v7 from http://lncrnakb.org.

#### NONCODE

NONCODE database integrated annotations from both literature searches and other public databases. The latest version, NONCODE version 6, is the single largest collection of lncRNAs, describing 96,422 lncRNA genes in humans. Each lncRNA gene in the NONCODE database had been assigned a unique NONCODE ID. We download the whole NONCODE v6 human data from http://www.noncode.org/datadownload/NONCODEv6_hg38.lncAndGene.bed.gz.

#### Integration across the lncRNA annotations

We kept only those lncRNA genes ranging in length from 200 nucleotides (nt) to 5 kilobases (kb). We limited the maximum length of a lncRNA gene to 5kb to control for the computational complexity^52^. Overlapping lncRNA genes between FANTOM and GENCODE using the Ensembl stable identifier were removed. We split each annotation file into individual files by chromosome with the start and end coordinates of the lncRNA genes. All duplicated lncRNAs between annotation files were removed by checking whether they have the same start and end coordinates. We then used the following intersection order based on experimental validation to merge the four lncRNA annotations: 1. GENCODE, 2. FANTOM5 CAT, 3. NONCODE and 4. lncRNAKB. Approximately 165k lncRNA genes were left for further analysis.

### LncRNA rare variant association test

#### lncRNA rare variant sets

We obtained the start and end genomic coordinates (human genome build GRCh38) of the lncRNA genomic regions from our previously curated lncRNA gene list. We then defined aggregation units by using all the rare variants (MAF <0.01) based on their genomic locations with respect to the start and end genomic coordinates of the lncRNA genes. We removed lncRNA rare variant sets that had less than two rare variants. For sensitivity analysis, we only aggregated exonic and splicing variants in lncRNA genes provided by GENCODE v29, for which is the default genome annotation employed by TOPMed consortium^34^.

#### STAAR unconditional analysis

We applied the STAAR (variant-set test for association using annotation information) framework to identify rare variants in the lncRNA variant sets that are associated with four quantitative lipid traits (LDL-C, HDL-C, TG and TC). STAAR is a scalable and powerful variant-set test that uses an omnibus multi-dimensional weighting scheme to incorporate both qualitative functional categories and multiple in silico variant annotation scores for genetic variants. STAAR accounts for population structure and relatedness and is scalable for analyzing large WGS studies of continuous and dichotomous traits by fitting linear and logistic mixed models^37,38^. To perform the STAAR unconditional analysis, we first fitted a STAAR null model using *fit_null_glmmkin()* function to account for sample relatedness with phenotypic data, covariates and (sparse) genetic relatedness matrix as input. For each of the four lipid phenotypes, we adjusted for age, age2, sex, study and PC1-PC11. We calculated the *P* value for each lncRNA rare variant set using STAAR-O, an omnibus test in the STAAR framework that combines *P* values from multiple annotation-weighted burden tests, SKAT and ACAT-V using the ACAT method. A total of 13 aggregated variant functional annotations were incorporated in STAAR-O, including three integrative scores (CADD^53^, LINSIGHT^54^ and FATHMM-XF^55^) and 10 annotation principal components (aPCs) (**Supplementary Table 5**)^38^. All analyses were performed using R packages *STAAR* (version 0.9.6) and *STAARpipeline* (version 0.9.6).

#### STAAR conditional analysis adjusting for known GLGC GWAS variants

We performed conditional analysis to identify lncRNA rare variant association independent of known lipid-associated variants. We obtained a list of 1,750 significant index variants (**Supplementary Table 6)** associated with one or more lipid levels from The Global Lipids Genetics Consortium (GLGC) latest lipid GWAS results^18,19,56^. The positions of SNV were lifted over to genome build 38. We adjusted for known lipid variants in a ±500 kb window beyond a variant set.

#### STAAR rare variant association test adjusting for nearby protein coding genes

The unconditional analysis showed that most lncRNA genes associated with lipids are near known monogenic lipid genes. We sought to perform conditional analyses adjusting lncRNA rare variant sets for nearby protein coding genes. The adjusted nearby protein coding genes can be divided into two categories: the closest protein coding genes and those nearby known lipid monogenic genes, including *ANGPTL8, APOA1, APOA5, APOB, APOC1, APOC3, APOE, CETP, LDLR, LPA, LPL, PCSK7, PCSK9, PLA2G15, TM6SF2*^19^. Our primary analysis was to adjust for only rare nonsynonymous variants (MAF < 1%) within nearby protein coding genes. We did two sensitivity analyses, one adjusted for rare synonymous variants (MAF < 1%) within nearby protein coding genes, and another adjusted for rare predicted loss-of-function (pLoF) variants (MAF < 1%) within nearby protein coding genes. For each participant, we created three burden scores separately by combining the minor allele counts of nonsynonymous, synonymous, and pLoF variants with a MAF < 1% carried within the closest gene and the nearby lipid monogenic genes in a 250kb window. We re-fitted null models similar to the unconditional analysis and added all the burden scores of the closest gene and the nearby lipid monogenic genes (if any) as additional covariates for each lipid phenotype. We then repeated the STAAR procedures to calculate the STAAR-O *P* values after adjusting for rare nonsynonymous, rare synonymous, and rare pLoF variants.

#### Effective number of independent tests

Although we removed redundant lncRNAs, the remaining lncRNAs can still have overlapping regions across different genome annotations. Therefore, we adopted a principal component analysis (PCA) based approach, the simple*M* method to calculate the effective number of independent tests^42^. For each chromosome, suppose we had tested K lncRNA rare variant set (lncRNA_1_, lncRNA_2_, …, lncRNA_K_) for N individuals (1, 2, …, N), we first found the minor allele counts of rare variants (MAF < 1%) carried by each individual within each lncRNA rare variant set that were tested by STAAR and constructed a NX K matrix. We then derived the pairwise lncRNA correlation matrix R_KxK_ that reflected the correlation structure among the tests from the constructed NX K matrix. We calculated the eigenvalues, {λ_*i*_:λ_l_ ≥ λ_l_ ≥ … ≥ λ_*K*_}, from the pairwise lncRNA correlation matrix R_KxK_. The effective number of tests (*M_eff_*) for each chromosome was estimated as 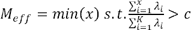, where c was a pre-defined parameter which was set to 0.95. We added up the effective number of tests (*M_eff_*) by each chromosome assuming independence between chromosomes. The Bonferroni correction formula was then used to calculate the adjusted significance level as 0.05/ *M_eff_* as used for unconditional analysis.

### LncRNA gene expression analysis

#### Framingham Heart Study (FHS) RNA-seq data

We utilized FHS RNA sequencing data to perform the association analyses of lncRNA expression levels with blood lipid traits. This study included 1505 participants from the FHS Third Generation cohort^41^. Blood samples for RNA seq were collected from Third Generation participants who attended the second examination cycle (2008–2011). Protocols for participant examinations and collection of genetic materials were approved by the Institutional Review Board at Boston Medical Center. All participants provided written, informed consent for genetic studies. All research was performed in accordance with relevant guidelines/regulations. The technical details for the blood draw and RNA sequencing can be found elsewhere^43^. For the association analyses (**Supplementary Text**), we first processed the RNASeq Data with following steps: 1. Sample QC by removing misidentified samples and sentinel control samples. 2. TMM normalization for the gene-level count data. 3. Filtering low expression transcripts. 4. Regressing the log2(TMM+1) on the technical covariates, and the resultant residuals were used to perform association analysis. We fitted a linear mixed effects model for the residuals of the TMM normalized log2 transformed counts data and the lipid phenotypes adjusting for predicted complete blood count (CBC), constructed surrogate variables (SVs), sex, age, and family structure as variance-covariance matrix.

## Genome build

All genome coordinates are given in the NCBI GRCh38/UCSC hg38 version of the human genome.

## Data Availability

Individual whole-genome sequence data for TOPMed and harmonized lipids at individual sample level are available through restricted access via the TOPMed dbGaP Exchange area. Summary level genotype data from TOPMed are available through the BRAVO browser (https://bravo.sph.umich.edu/). The UK Biobank (UKB) whole-genome sequence data can be accessed through UKB Research Analysis Platform (RAP), through the UKB approval system (https://www.ukbiobank.ac.uk). The dbGaP accessions for TOPMed cohorts are as follows: Old Order Amish (Amish) phs000956 and phs00039;Atherosclerosis Risk in Communities study (ARIC) phs001211 and phs000280; Mt Sinai BioMe Biobank (BioMe) phs001644 and phs000925; Coronary Artery Risk Development in Young Adults (CARDIA) phs001612 and phs000285; Cleveland Family Study (CFS) phs000954 and phs000284; Cardiovascular Health Study (CHS) phs001368 and phs000287; Diabetes Heart Study (DHS) phs001412 and phs001012; Framingham Heart Study (FHS) phs000974 and phs000007; Genetic Studies of Atherosclerosis Risk (GeneSTAR) phs001218 and phs000375; Genetic Epidemiology Network of Arteriopathy (GENOA) phs001345 and phs001238; Genetic Epidemiology Network of Salt Sensitivity (GenSalt) phs001217 and phs000784; Genetics of Lipid-Lowering Drugs and Diet Network (GOLDN) phs001359 and phs000741; Hispanic Community Health Study - Study of Latinos (HCHS_SOL) phs001395 and phs000810; Hypertension Genetic Epidemiology Network and Genetic Epidemiology Network of Arteriopathy (HyperGEN) phs001293 and phs001293; Jackson Heart Study (JHS) phs000964 and phs000286; Multi-Ethnic Study of Atherosclerosis (MESA) phs001416 and phs000209; Massachusetts General Hospital Atrial Fibrillation Study (MGH_AF) phs001062 and phs001001; San Antonio Family Study (SAFS) phs001215 and phs000462; Samoan Adiposity Study (SAS) phs000972 and phs000914; Taiwan Study of Hypertension using Rare Variants (THRV) phs001387 and phs001387; Women's Health Initiative (WHI) phs001237 and phs000200.

## Acknowledgements

Whole genome sequencing (WGS) for the Trans-Omics in Precision Medicine (TOPMed) program was supported by the National Heart, Lung and Blood Institute (NHLBI). G.M.P. is supported by NIH grants R01HL142711 and R01HL127564. P.N. is supported by grants from the National Heart, Lung, and Blood Institute (R01HL142711, R01HL148050, R01HL151283, R01HL148565, R01HL135242, R01HL151152), Fondation Leducq (TNE-18CVD04), and Massachusetts General Hospital (Paul and Phyllis Fireman Endowed Chair in Vascular Medicine). X.Lin is supported by grants R35-CA197449, U19-CA203654, R01-HL113338, and U01-HG009088. We like to acknowledge all the grants that supported this study, R01 HL121007, U01 HL072515, R01 AG18728, X01HL134588, HL 046389, HL113338, and 1R35HL135818, K01 HL135405, R03 HL154284, U01HL072507, R01HL087263, R01HL090682, P01HL045522, R01MH078143, R01MH078111, R01MH083824, U01DK085524, R01HL113323, R01HL093093, R01HL140570, R01HL142711, R01HL127564, R01HL148050, R01HL148565, HL105756, and Leducq TNE-18CVD04. The views expressed in this manuscript are those of the authors and do not necessarily represent the views of the National Heart, Lung, and Blood Institute; the National Institutes of Health; or the U.S. Department of Health and Human Services. We gratefully acknowledge the studies and participants who provided biological samples and data for TOPMed and UK Biobank. The full study specific acknowledgements and NHLBI TOPMed Fellowship acknowledgement are detailed in **Supplementary Text**.

## Author contributions

Y.W., P.N., and G.M.P. designed the study. Y.W. carried out all the primary analysis with critical inputs from P.N. and G.M.P. M.S.S carried out the replication analysis. Y.W. and J.A.H carried out the secondary analysis. Y.W., M.S.S., X.Li, Z.L., A.K.D, J.C.B., J.B., E.B., D.W.B., B.E.C., J.C.C., A.P.C., Y.C., J.E.C., P.S.D., S.K.D., P.T.E., J.S.F., M.F., B.I.F., S. Gabriel, S.Germer, R.A.G., X.G., J.H., N.H., B.H., L.H., M.R.I., R.J., R.C.K., S.LR.K., T.N.K., R.K., C.K., B.G.K., D.Levy, C.Li, C.Liu, D.Lloyd-Jone, R.JF.L., M.C.M., L.W.M., R.A.M., R.L.M., B.D.M., M.E.M., A.C.M., J.M.M., T.N., J.R.O., N.D.P., M.H.P., B.M.P., L.M.E., D.C.R., S.R., A.P.R., S.S.R., M.R., W.H-H.S., J.A.S., A.S., H.K.T., M.Y.T., K.A.V., Z.W., L.R.Y., W.Z., J.I.R., X.Lin., P.N., and G.M.P. acquired, analyzed or interpreted data. G.M.P. and P.N. and NHLBI TOPMed Lipids Working Group provided administrative, technical, or material support. Y.W. and G.M.P. wrote the first draft of the manuscript and revised it according to suggestions by the coauthors. All authors critically reviewed the manuscript, suggested revisions as needed and approved the final version.

## Data availability

Individual whole-genome sequence data for TOPMed and harmonized lipids at individual sample level are available through restricted access via the TOPMed dbGaP Exchange area. Summary level genotype data from TOPMed are available through the BRAVO browser (https://bravo.sph.umich.edu/). The UK Biobank (UKB) whole-genome sequence data can be accessed through UKB Research Analysis Platform (RAP), through the UKB approval system (https://www.ukbiobank.ac.uk). The dbGaP accessions for TOPMed cohorts are as follows: Old Order Amish (Amish) *phs000956 and phs00039;*Atherosclerosis Risk in Communities study (ARIC) *phs001211 and phs000280;* Mt Sinai BioMe Biobank (BioMe) *phs001644 and phs000925;* Coronary Artery Risk Development in Young Adults (CARDIA) phs001612 and phs000285; Cleveland Family Study (CFS) *phs000954 and phs000284;* Cardiovascular Health Study (CHS) *phs001368 and phs000287;* Diabetes Heart Study (DHS) *phs001412 and phs001012;* Framingham Heart Study (FHS) *phs000974 and phs000007;* Genetic Studies of Atherosclerosis Risk (GeneSTAR) *phs001218 and phs000375;* Genetic Epidemiology Network of Arteriopathy (GENOA) *phs001345 and phs001238;* Genetic Epidemiology Network of Salt Sensitivity (GenSalt) *phs001217 and phs000784;* Genetics of Lipid-Lowering Drugs and Diet Network (GOLDN) *phs001359 and phs000741;* Hispanic Community Health Study - Study of Latinos (HCHS_SOL) *phs001395 and phs000810;* Hypertension Genetic Epidemiology Network and Genetic Epidemiology Network of Arteriopathy (HyperGEN) *phs001293 and phs001293;* Jackson Heart Study (JHS) *phs000964 and phs000286;* Multi-Ethnic Study of Atherosclerosis (MESA) *phs001416 and phs000209;* Massachusetts General Hospital Atrial Fibrillation Study (MGH_AF) *phs001062 and phs001001;* San Antonio Family Study (SAFS) *phs001215 and phs000462;* Samoan Adiposity Study (SAS) *phs000972 and phs000914;* Taiwan Study of Hypertension using Rare Variants (THRV) *phs001387 and phs001387;* Women’s Health Initiative (WHI) *phs001237 and phs000200*.

## Code availability

R code for implementing the analysis is available at the public GitHub Repository https://github.com/kyleyxw/lncRNA-paper. STAAR is implemented as an open-source R package available at https://github.com/xihaoli/STAAR. STAARpipeline is implemented as an open-source R package available at https://github.com/xihaoli/STAARpipeline.

