## Supplementary Text for "Rare variants in long non-coding RNAs are associated with blood lipid levels in the TOPMed Whole Genome Sequencing Study"

[Supplementary Figure 2. Results for the 61 lncRNA-lipid associations that remained significant (STAAR-O P value < 6.0e-04) in the conditional analysis adjusting for known lipid-associated GWAS variants.	20](#_Toc138759631)

[Supplementary Figure 2. Results for the 61 lncRNA-lipid associations that remained significant (STAAR-O P value < 6.0e-04) in the conditional analysis adjusting for known lipid-associated GWAS variants.	24](#_Toc138759642)

### Supplementary Text: TOPMed Cohorts description

#### Old Order Amish (Amish, n=1,083):

*TOPMed dbGaP accession#: phs000956, Parent dbGaP accession#: phs000391, Sequencing Center: Broad Institute of MIT and Harvard.*

The Old Order Amish (OOA) population of Lancaster County, PA immigrated to the Colonies from Western Europe in the early 1700’s. Investigators at University of Maryland School of Medicine have been studying the genetic determinants of cardiometabolic health in this population since 1993. To date, over 7,000 Amish adults have participated in one or more of our studies. The Heredity and Phenotype Intervention (HAPI) Heart Study was initiated in 2002 and chose to study the OOA to test the genetic effects of complex phenotypes from this genetically homogeneous population^1^. The Amish Research Group includes investigators with a diverse range of interests in population and basic science and in clinical and translational research. Whole genome sequencing (WGS) for the Trans-Omics in Precision Medicine (TOPMed) program was supported by the National Heart, Lung and Blood Institute (NHLBI). The Amish studies were supported by NIH grants R01 AG18728, U01 HL072515, R01 HL088119, R01 HL121007, and P30 DK072488.

#### Atherosclerosis Risk in Communities study (ARIC, n=8,016):

*TOPMed dbGaP accession#: phs001211, Parent dbGaP accession#: phs000280, Sequencing Center: Baylor College of Medicine Human Genome Sequencing Center and Broad Institute of MIT and Harvard.*

The ARIC study is a population-based prospective cohort study of cardiovascular disease sponsored by the National Heart, Lung, and Blood Institute (NHLBI). ARIC included 15,792 individuals, predominantly European American and African American, aged 45-64 years at baseline (1987-89), chosen by probability sampling from four US communities. Cohort members completed three additional triennial follow-up examinations, a fifth exam in 2011-2013, a sixth exam in 2016-2017, a seventh exam in 2018-2019, and an eighth exam in 2020. The ARIC study has been described in detail previously^2^.

Whole genome sequencing (WGS) for the Trans-Omics in Precision Medicine (TOPMed) program was supported by the National Heart, Lung and Blood Institute (NHLBI). WGS for “NHLBI TOPMed: Atherosclerosis Risk in Communities (ARIC)” (phs001211) was performed at the Baylor College of Medicine Human Genome Sequencing Center (HHSN268201500015C and 3U54HG003273-12S2) and the Broad Institute for MIT and Harvard (3R01HL092577-06S1). Centralized read mapping and genotype calling, along with variant quality metrics and filtering were provided by the TOPMed Informatics Research Center (3R01HL-117626-02S1). Phenotype harmonization, data management, sample-identity QC, and general study coordination, were provided by the TOPMed Data Coordinating Center (3R01HL-120393-02S1). We gratefully acknowledge the studies and participants who provided biological samples and data for TOPMed.

The Genome Sequencing Program (GSP) was funded by the National Human Genome Research Institute (NHGRI), the National Heart, Lung, and Blood Institute (NHLBI), and the National Eye Institute (NEI). The GSP Coordinating Center (U24 HG008956) contributed to cross-program scientific initiatives and provided logistical and general study coordination. The Centers for Common Disease Genomics (CCDG) program was supported by NHGRI and NHLBI, and whole genome sequencing was performed at the Baylor College of Medicine Human Genome Sequencing Center (UM1 HG008898 and R01HL059367).

The Atherosclerosis Risk in Communities study has been funded in whole or in part with Federal funds from the National Heart, Lung, and Blood Institute, National Institutes of Health, Department of Health and Human Services, under Contract numbers 75N92022D00001, 75N92022D00002, 75N92022D00003, 75N92022D00004, 75N92022D00005. The authors thank the staff and participants of the ARIC study for their important contributions.

#### Mt Sinai BioMe Biobank (BioMe, n=9,848):

*TOPMed dbGaP accession#: phs001644, Parent dbGaP accession#: phs000925, Sequencing Center: Baylor College of Medicine Human Genome Sequencing Center, Northwest Genomics Center*

The Institute for Personalized Medicine at the Icahn School of Medicine at Mount Sinai is leading the movement toward diagnosis and classification of disease according to the patient’s molecular profile. BioMe is the major effort towards this goal, BioMe, an electronic medical record-linked biobank that enables researchers to conduct genetic, epidemiologic, molecular, and genomic studies rapidly and efficiently on large collections of research specimens linked with medical information. BioMed cohort is composed of White, Black, Hispanic and Asian populations. The Mount Sinai Medical Center services diverse local communities of upper Manhattan, including Central Harlem (86% African American), East Harlem (88% Hispanic Latino), and Upper East Side (88% Caucasian/white) with broad health disparities^3^. Whole genome sequencing (WGS) for the Trans-Omics in Precision Medicine (TOPMed) program was supported by the National Heart, Lung and Blood Institute (NHLBI). The Mount Sinai BioMe Biobank has been supported by The Andrea and Charles Bronfman Philanthropies and in part by Federal funds from the NHLBI and NHGRI (U01HG00638001; U01HG007417; X01HL134588).

#### Coronary Artery Risk Development in Young Adults (CARDIA, n=3,056):

TOPMed dbGaP accession#: phs001612, Parent dbGaP accession#: phs000285, *Sequencing Center: Baylor College of Medicine Human Genome Sequencing Center.*

The Coronary Artery Risk Development in Young Adults (CARDIA) Study is a study examining the development and determinants of clinical and subclinical cardiovascular disease and their risk factors. It began in 1985-6 with a group of 5115 black and white men and women aged 18-30 years^4^. The participants were selected so that there would be approximately the same number of people in subgroups of race, gender, education (high school or less and more than high school) and age (18-24 and 25-30) in each of 4 centers: Birmingham, AL; Chicago, IL; Minneapolis, MN; and Oakland, CA. These same participants were asked to participate in follow-up examinations during 1987-1988 (Year 2), 1990-1991 (Year 5), 1992-1993 (Year 7), 1995-1996 (Year 10), 2000-2001 (Year 15), 2005-2006 (Year 20), 2010-2011 (Year 25), and 2015-2016 (Year 30), 2020-2023 (Year 35). The CARDIA cohort is composed of White and Black populations. Whole genome sequencing (WGS) for the Trans-Omics in Precision Medicine (TOPMed) program was supported by the National Heart, Lung and Blood Institute (NHLBI). The Coronary Artery Risk Development in Young Adults Study (CARDIA) is conducted and supported by the National Heart, Lung, and Blood Institute (NHLBI) in collaboration with the University of Alabama at Birmingham (HHSN268201800005I & HHSN268201800007I), Northwestern University (HHSN268201800003I), University of Minnesota (HHSN268201800006I), and Kaiser Foundation Research Institute (HHSN268201800004I). CARDIA was also partially supported by the Intramural Research Program of the National Institute on Aging (NIA) and an intra‐agency agreement between NIA and NHLBI (AG0005).

#### Cleveland Family Study (CFS, n=579):

*TOPMed dbGaP accession#: phs000954, Parent dbGaP accession#: phs000284, Sequencing Center:* *Northwest Genomics Center.*

The CFS is a family-based longitudinal study that includes participants with laboratory diagnosed sleep apnea, their family members and neighborhood control families followed between 1990 and 2006^5^. After an overnight fast, blood was collected which was assayed for lipid levels at the University of Vermont Laboratory for Clinical Biochemistry Research. Lipids (triglycerides, HDL cholesterol) from fasted blood serum were measured by enzymatic methods. The CFS cohort is composed of White and Black populations. Whole genome sequencing (WGS) for the Trans-Omics in Precision Medicine (TOPMed) program was supported by the National Heart, Lung and Blood Institute (NHLBI). CFS is supported by grants from the NHLBI (R01 HL46380, R01 HL113338, and 1R35HL135818).

#### Cardiovascular Health Study (CHS, n=3,456):

*TOPMed dbGaP accession#: phs001368, Parent dbGaP accession#: phs000287, Sequencing Center: Baylor College of Medicine Human Genome Sequencing Center.*

The Cardiovascular Health Study (CHS) is an NHLBI-funded observational study of risk factors for cardiovascular disease in adults 65 years or older. Starting in 1989, and continuing through 1999, participants underwent annual extensive clinical examinations. Measurements included traditional risk factors such as blood pressure and lipids. Additionally, measures of subclinical disease, including echocardiography of the heart, carotid ultrasound, and cranial magnetic-resonance imaging (MRI) were determined. At six-month intervals between clinic visits, and once clinic visits ended, participants were contacted by phone to ascertain hospitalizations and health status. The main outcomes are coronary heart disease (CHD), angina, heart failure (HF), stroke, transient ischemic attack (TIA), claudication, and mortality^6^. Participants continue to be contacted by phone every 6 months. Participants from four counties were included in the study: Forsyth County, North Carolina; Sacramento County, California; Washington County, Maryland; and Pittsburgh, Pennsylvania. CHS cohort is composed of White and Black populations. Whole genome sequencing (WGS) for the Trans-Omics in Precision Medicine (TOPMed) program was supported by the National Heart, Lung and Blood Institute (NHLBI). The CHS research was supported by NHLBI contracts 75N92021D00006, HHSN268201200036C, HHSN268200800007C, HHSN268201800001C, N01HC55222, N01HC85079, N01HC85080, N01HC85081, N01HC85082, N01HC85083, N01HC85086; and NHLBI grants U01HL080295, R01HL087652, R01HL105756, R01HL103612, R01HL120393, R01HL130114, and R01 HL059367, with additional contribution from the National Institute of Neurological Disorders and Stroke (NINDS). Additional support was provided through R01AG023629 from the National Institute on Aging (NIA).

#### Diabetes Heart Study (DHS, n=365):

*TOPMed dbGaP accession#: phs001412, Parent dbGaP accession#: phs001012, Sequencing Center: Broad Institute of MIT and Harvard*

The Diabetes Heart Study (DHS) is a family-based study enriched for type 2 diabetes (T2D). The cohort was recruited between 1998 and 2006. Participants were extensively phenotyped for measures of subclinical CVD and other known CVD risk factors^7^. Primary outcomes were quantified burden of vascular calcified plaque in the coronary artery, carotid artery, and abdominal aorta all determined from non-contrast computed tomography scans. Whole genome sequencing (WGS) for the Trans-Omics in Precision Medicine (TOPMed) program was supported by the National Heart, Lung and Blood Institute (NHLBI). The DHS research was supported by R01 HL92301, R01 HL67348, R01 NS058700, R01 AR48797, R01 DK071891, R01 AG058921, the General Clinical Research Center of the Wake Forest University School of Medicine (M01 RR07122, F32 HL085989), the American Diabetes Association, and a pilot grant from the Claude Pepper Older Americans Independence Center of Wake Forest University Health Sciences (P60 AG10484).

#### Framingham Heart Study (FHS, n=3,992):

*TOPMed dbGaP accession#: phs000974, Parent dbGaP accession#: phs000007, Sequencing Center: Broad Institute of MIT and Harvard*

The Framingham Heart Study (FHS) is a prospective cohort study of 3 generations of subjects who have been followed up to 65 years to evaluate risk factors for cardiovascular disease.^8–11^ Its large sample of ~15,000 men and women who have been extensively phenotyped with repeated examinations make it ideal for the study of genetic associations with cardiovascular disease risk factors and outcomes. DNA samples have been collected and immortalized since the mid-1990s and are available on ~8000 study participants in 1037 families. These samples have been used for collection of GWAS array data and exome chip data in nearly all with DNA samples, and for targeted sequencing, deep exome sequencing and light coverage whole genome sequencing in limited numbers. Additionally, mRNA and miRNA expression data, DNA methylation data, metabolomics and other 'omics data are available on a sizable portion of study participants. This project on the focuses on deep whole genome sequencing (mean 30X coverage) in 3,992 subjects with lipid data available.

FHS acknowledges the support of contracts NO1-HC-25195 and HHSN268201500001I from the National Heart, Lung and Blood Institute and grants supplement R01 HL092577-06S1 for this research. WGS for “NHLBI TOPMed: Whole Genome Sequencing and Related Phenotypes in the Framingham Heart Study” (phs000974) was performed at the Broad Institute of MIT and Harvard (HHSN268201500014C, 3R01HL092577-06S1, and 3U54HG003067-12S2). We also acknowledge the dedication of the FHS study participants without whom this research would not be possible.

#### Genetic Studies of Atherosclerosis Risk (GeneSTAR, n=1,757):

*TOPMed dbGaP accession#: phs001218, Parent dbGaP accession#: phs000375, Sequencing Center: Broad Institute of MIT and Harvard, Illumina Genomic Services, PSOMAGEN (formerly Macrogen).*

GeneSTAR is a family-based study in initially healthy brothers and sisters, and offspring of people with early-onset coronary disease. The goal is to discover and amplify mechanisms of stroke and coronary heart disease. In 1982 GeneSTAR was created to study patterns of coronary heart disease and related risk factors in families with early-onset coronary disease, identified from10 Baltimore area Hospitals. Extensive additional cardiovascular testing and risk assessment was done at baseline and serially^12,13^. Follow-up was carried out to determine incident cardiovascular disease, stroke, peripheral arterial disease, diabetes, cancer, and related comorbidities, from 5 to 30 years after study entry. Whole genome sequencing (WGS) for the Trans-Omics in Precision Medicine (TOPMed) program was supported by the National Heart, Lung and Blood Institute (NHLBI). GeneSTAR was supported by grants from the National Institutes of Health/National Heart, Lung, and Blood Institute (U01 HL72518, HL087698, HL49762, HL58625, HL071025, HL112064), the National Institutes of Health/National Institute of Nursing Research (NR0224103), and by a grant from the National Institutes of Health/National Center for Research Resources (M01-RR000052) to the Johns Hopkins General Clinical Research Center.

#### Genetic Epidemiology Network of Arteriopathy (GENOA, n=1,046):

*TOPMed dbGaP accession#: phs001345, Parent dbGaP accession#: phs001238, Sequencing Center: Broad Institute of MIT and Harvard, McDonnell Genome Institute (MGI) at Washington University.*

The Genetic Epidemiology Network of Arteriopathy (GENOA) is one of four networks in the NHLBI Family-Blood Pressure Program (FBPP)^14^. GENOA's long-term objective is to elucidate the genetics of target organ complications of hypertension, including both atherosclerotic and arteriolosclerotic complications involving the heart, brain, kidneys, and peripheral arteries^15^. The longitudinal GENOA Study recruited European-American and African-American sibships with at least 2 individuals with clinically diagnosed essential hypertension before age 60 years. All other members of the sibship were invited to participate regardless of their hypertension status. Participants were diagnosed with hypertension if they had either 1) a previous clinical diagnosis of hypertension by a physician with current anti-hypertensive treatment, or 2) an average systolic blood pressure ≥ 140 mm Hg or diastolic blood pressure ≥ 90 mm Hg based on the second and third readings at the time of their clinic visit. Only participants of the African-American Cohort were sequenced through TOPMed. Whole genome sequencing (WGS) for the Trans-Omics in Precision Medicine (TOPMed) program was supported by the National Heart, Lung and Blood Institute (NHLBI). Support for GENOA was provided by the National Heart, Lung and Blood Institute (HL054457, HL054464, HL054481, and HL087660) of the National Institutes of Health.

#### Genetic Epidemiology Network of Salt Sensitivity (GenSalt, n=1,772):

*TOPMed dbGaP accession#: phs001217, Parent dbGaP accession#: phs000784, Sequencing Center: Broad Institute of MIT and Harvard.*

GenSalt utilizes a family feeding-study design. Each family is ascertained through a proband with untreated prehypertension or stage-1 hypertension in rural China. Medical history, lifestyle risk factors, and cold pressor tests are obtained at baseline visits while BP, weight, blood and urine specimens are collected at baseline and follow-up visits^16^. The dietary intervention includes a 7-day low sodium-feeding (51.3 mmol/day), a 7-day high sodium-feeding (307.8 mmol/day), and a 7-day high sodium-feeding with an oral potassium supplementation (60 mmol/day). Whole genome sequencing (WGS) for the Trans-Omics in Precision Medicine (TOPMed) program was supported by the National Heart, Lung and Blood Institute (NHLBI). GenSalt was supported by research grants (U01HL072507, R01HL087263, and R01HL090682) from the National Heart, Lung and Blood Institute, National Institutes of Health, Bethesda, MD.

#### Genetics of Lipid-Lowering Drugs and Diet Network (GOLDN, n=926):

*TOPMed dbGaP accession#: phs001359, Parent dbGaP accession#: phs000741, Sequencing Center: McDonnell Genome Institute (MGI) at Washington University.*

GOLDN is a family-based study of European descent individuals recruited in Minneapolis and Salt Lake City (two of the NHLBI Family Heart Study sites). Contributions of genes, shared and individual environments, and behaviors to variations in risk factors, preclinical atherosclerosis, and CHD were estimated in the study^17^. It aims to uncover genetic predictors of variability in lipid phenotypes, which include both fasting and postprandial lipids quantified using traditional methods, NMR, and high throughput lipidomics. Whole genome sequencing (WGS) for the Trans-Omics in Precision Medicine (TOPMed) program was supported by the National Heart, Lung and Blood Institute (NHLBI). GOLDN biospecimens, baseline phenotype data, and intervention phenotype data were collected with funding from National Heart, Lung and Blood Institute (NHLBI) grant U01 HL072524.

#### Hispanic Community Health Study - Study of Latinos (HCHS_SOL, n=7714):

*TOPMed dbGaP accession#: phs001395, Parent dbGaP accession#: phs000810, Sequencing Center: Baylor College of Medicine Human Genome Sequencing Center.*

The Hispanic Community Health Study (HCHS)/Study of Latinos (SOL) is a multicenter, community-based cohort study of Hispanic/Latino adults in the United States. The main goal of the study is to identify risk factors which could either be protective or harmful to the Hispanic community^18,19^. A total of 16,000 Hispanic/Latino individuals of age 18-74 were recruited. Participants are recruited in community areas surrounding four field centers in the Bronx, Chicago, Miami, and San Diego. Whole genome sequencing (WGS) for the Trans-Omics in Precision Medicine (TOPMed) program was supported by the National Heart, Lung and Blood Institute (NHLBI). The Hispanic Community Health Study/Study of Latinos was carried out as a collaborative study supported by contracts from the National Heart, Lung, and Blood Institute (NHLBI) to the University of North Carolina (N01-HC65233), University of Miami (N01-HC65234), Albert Einstein College of Medicine (N01-HC65235), Northwestern University (N01-HC65236), and San Diego State University (N01-HC65237).

#### Hypertension Genetic Epidemiology Network and Genetic Epidemiology Network of Arteriopathy (HyperGEN, n=1,853):

*TOPMed dbGaP accession#: phs001293, Parent dbGaP accession#: phs001293, Sequencing Center: McDonnell Genome Institute (MGI) at Washington University.*

The Hypertension Genetic Epidemiology Network Study (HyperGEN) - Genetics of Left Ventricular (LV) Hypertrophy is a familial study aimed to understand genetic risk factors for LV hypertrophy by conducting genetic studies of continuous traits from echocardiography exams^20,21^. As part of HyperGEN study, four field centers recruited African American and white hypertensive siblings, aged 23 to 87 years. The current study includes only African Americans from HyperGEN. Data from detailed clinical exams as well as genotyping data for linkage studies, candidate gene studies and GWAS have been collected and is shared between HyperGEN and the ancillary HyperGEN - Genetics of LV Hypertrophy study. Whole genome sequencing (WGS) for the Trans-Omics in Precision Medicine (TOPMed) program was supported by the National Heart, Lung and Blood Institute (NHLBI). The HyperGEN Study is part of the National Heart, Lung, and Blood Institute (NHLBI) Family Blood Pressure Program; collection of the data represented here was supported by grants U01 HL054472 (MN Lab), U01 HL054473 (DCC), U01 HL054495 (AL FC), and U01 HL054509 (NC FC). The HyperGEN: Genetics of Left Ventricular Hypertrophy Study was supported by NHLBI grant R01 HL055673 with whole-genome sequencing made possible by supplement -18S1.

#### Jackson Heart Study (JHS, n=2,847):

*TOPMed dbGaP accession#: phs000964, Parent dbGaP accession#: phs000286, Sequencing Center: McDonnell Genome Institute (MGI) at Washington University.*

The JHS is a large, community-based, observational study of African American adults residing in the Jackson, Mississippi metropolitan statistical area (MSA). Four subsamples of participants (random, volunteer, ARIC (continuing from Atherosclerosis Risk in Communities study), and family) were included ^22–24^. Recruitment was limited to persons 35-84 years old except in the family cohort, where those 21 years old and above were eligible. Participants provided extensive medical and social history, had an array of physical and biochemical measurements and diagnostic procedures, and provided genomic DNA. Whole genome sequencing (WGS) for the Trans-Omics in Precision Medicine (TOPMed) program was supported by the National Heart, Lung and Blood Institute (NHLBI). The Jackson Heart Study (JHS) is supported and conducted in collaboration with Jackson State University (HHSN268201800013I), Tougaloo College (HHSN268201800014I), the Mississippi State Department of Health (HHSN268201800015I) and the University of Mississippi Medical Center (HHSN268201800010I, HHSN268201800011I and HHSN268201800012I) contracts from the National Heart, Lung, and Blood Institute (NHLBI) and the National Institute on Minority Health and Health Disparities (NIMHD).The authors also wish to thank the staffs and participants of the JHS. The views expressed in this manuscript are those of the authors and do not necessarily represent the views of the National Heart, Lung, and Blood Institute; the National Institutes of Health; or the U.S. Department of Health and Human Services.

#### Multi-Ethnic Study of Atherosclerosis (MESA, n=5,290):

*TOPMed dbGaP accession#: phs001416, Parent dbGaP accession#: phs000209,*

*Sequencing Center: Broad Institute of MIT and Harvard.*

The Multi-Ethnic Study of Atherosclerosis (MESA) is a study of the characteristics of subclinical cardiovascular disease (disease detected non-invasively before it has produced clinical signs and symptoms) and the risk factors that predict progression to clinically overt cardiovascular disease or progression of the subclinical disease. MESA researchers study a diverse, population-based sample of 6,814 asymptomatic men and women aged 45-84 from six field centers across the United States. Approximately 38 % of the recruited participants are white, 28 % African American, 22 % Hispanic, and 12 % Asian, predominantly of Chinese descent^25^. Six exams have been completed since 2000. Participants are contacted every 9 to 12 months throughout the study to assess clinical morbidity and mortality. The final 18 months of the study will be dedicated to close out and data analysis and publication. Baseline measurements will include measurement of coronary calcium using computed tomography; measurement of ventricular mass and function using cardiac magnetic resonance imaging; measurement of flow-mediated brachial artery endothelial vasodilation, carotid intimal-medial wall thickness, and distensibility of the carotid arteries using ultrasonography; measurement of peripheral vascular disease using ankle and brachial blood pressures; electrocardiography; and assessments of microalbuminuria, standard CVD risk factors, sociodemographic factors, life habits, and psychosocial factors.

Centralized read mapping and genotype calling, along with variant quality metrics and filtering were provided by the TOPMed Informatics Research Center (3R01HL-117626-02S1). Phenotype harmonization, data management, sample-identity QC, and general study coordination, were provided by the TOPMed Data Coordinating Center (3R01HL-120393-02S1). MESA and the MESA SHARe project are conducted and supported by the National Heart, Lung, and Blood Institute (NHLBI) in collaboration with MESA investigators. Whole genome sequencing (WGS) for the Trans-Omics in Precision Medicine (TOPMed) program was supported by the National Heart, Lung and Blood Institute (NHLBI). WGS for “NHLBI TOPMed: Multi-Ethnic Study of Atherosclerosis (MESA)” (phs001416.v1.p1) was performed at the Broad Institute of MIT and Harvard (3U54HG003067-13S1). Centralized read mapping and genotype calling, along with variant quality metrics and filtering were provided by the TOPMed Informatics Research Center (3R01HL-117626-02S1). Phenotype harmonization, data management, sample-identity QC, and general study coordination, were provided by the TOPMed Data Coordinating Center (3R01HL-120393-02S1), and TOPMed MESA Multi-Omics (HHSN2682015000031/HSN26800004). The MESA projects are conducted and supported by the National Heart, Lung, and Blood Institute (NHLBI) in collaboration with MESA investigators. Support for the Multi-Ethnic Study of Atherosclerosis (MESA) projects are conducted and supported by the National Heart, Lung, and Blood Institute (NHLBI) in collaboration with MESA investigators. Support for MESA is provided by contracts 75N92020D00001, HHSN268201500003I, N01-HC-95159, 75N92020D00005, N01-HC-95160, 75N92020D00002, N01-HC-95161, 75N92020D00003, N01-HC-95162, 75N92020D00006, N01-HC-95163, 75N92020D00004, N01-HC-95164, 75N92020D00007, N01-HC-95165, N01-HC-95166, N01-HC-95167, N01-HC-95168, N01-HC-95169, UL1-TR-000040, UL1-TR-001079, UL1-TR-001420, UL1TR001881, DK063491, and R01HL105756. The authors thank the other investigators, the staff, and the participants of the MESA study for their valuable contributions. A full list of participating MESA investigators and institutes can be found at [http://www.mesa-nhlbi.org](http://www.mesa-nhlbi.org/).

#### Massachusetts General Hospital Atrial Fibrillation Study (MGH_AF, n=683):

*TOPMed dbGaP accession#: phs001062, Parent dbGaP accession#: phs001001*

*Sequencing Center: Broad Institute of MIT and Harvard.*

The MGH-AF study was initiated to study the effects of atrial fibrillations by comparing unaffected and affected family members^26,27^. The participants provide details on past medical history, AF treatment and family history. An electrocardiogram is performed; the results of an echocardiogram are obtained along with blood samples. For the TOPMed whole genome sequencing project only early-onset atrial fibrillation cases were sequenced. Early-onset atrial fibrillation was defined as an age of onset prior to 66 years of age. Whole genome sequencing (WGS) for the Trans-Omics in Precision Medicine (TOPMed) program was supported by the National Heart, Lung and Blood Institute (NHLBI). The MGH AF Study was supported by grants to Dr. Ellinor from the Fondation Leducq (14CVD01), the National Institutes of Health to Dr. Ellinor (1RO1HL092577, R01HL128914, K24HL105780) and Dr. Lubitz (1R01HL139731) and by grants from the American Heart Association to Dr. Ellinor (18SFRN34110082) and to Dr. Lubitz (18SFRN34250007).

#### San Antonio Family Study (SAFS, n=619):

*TOPMed dbGaP accession#: phs001215, Parent dbGaP accession#: phs000462, Sequencing Center: Illumina Genomic Services.*

The SAFS began in 1991, and included 1,431 individuals in 42 extended families at baseline. Probands were 40- to 60-year-old low-income Mexican Americans selected at random without regard to presence or absence of disease, almost exclusively from Mexican American census tracts in San Antonio, Texas. The major objectives of this study are to identify low frequency or rare variants in and around known common variant signals for CVD, as well as to find novel low frequency or rare variants influencing susceptibility to CVD^28^. Whole genome sequencing (WGS) for the Trans-Omics in Precision Medicine (TOPMed) program was supported by the National Heart, Lung and Blood Institute (NHLBI). Collection of the San Antonio Family Study data was supported in part by National Institutes of Health (NIH) grants R01 HL045522, MH078143, MH078111 and MH083824; and whole genome sequencing of SAFS subjects was supported by U01 DK085524 and R01 HL113323. We are very grateful to the participants of the San Antonio Family Study for their continued involvement in our research programs.

#### Samoan Adiposity Study (Samoan, n=1,182):

*TOPMed dbGaP accession#: phs000972, Parent dbGaP accession#: phs000914,*

*Sequencing Center: New York Genome Center and McDonnell Genome Institute (MGI) at Washington University.*

The main aim of the Samoan Adiposity Study is to identify risk factors for obesity and cardiometabolic phenotypes among the Samoan population. The recruitment and measurement of the study participants took place from February to July 2010 and thirty-three villages were included in the study^29,30^. The participants reside throughout the independent nation of Samoa, which is experiencing economic development and the nutrition transition. Genotyping was performed with the Affymetrix Genome-Wide Human SNP 6.0 Array using a panel of approximately 900,000 SNPs. Anthropometric, fasting blood biomarkers and detailed dietary, physical activity, health and socio-demographic variables were collected. Whole genome sequencing (WGS) for the Trans-Omics in Precision Medicine (TOPMed) program was supported by the National Heart, Lung and Blood Institute (NHLBI). Data collection was funded by NIH grant R01-HL093093. We thank the Samoan participants of the study and local village authorities. We acknowledge the support of the Samoan Ministry of Health and the Samoa Bureau of Statistics for their support of this research.

#### Taiwan Study of Hypertension using Rare Variants (THRV, n=1,982):

*TOPMed dbGaP accession#: phs001387, Parent dbGaP accession#: phs001387, Sequencing Center: Baylor College of Medicine Human Genome Sequencing Center.*

The THRV-TOPMed study consists of three cohorts: The SAPPHIRe Family cohort, TSGH (Tri-Service General Hospital, a hospital-based cohort), and TCVGH (Taichung Veterans General Hospital, another hospital-based cohort), all based in Taiwan^31,32^. 1,271 subjects were previously recruited as part of the NHLBI-sponsored SAPPHIRe Network (which is part of the Family Blood Pressure Program, FBPP). THRV is a collaborative study between Washington University in St. Louis, LA BioMed at Harbor UCLA, University of Texas in Houston, Taichung Veterans General Hospital, Taipei Veterans General Hospital, Tri-Service General Hospital, National Health Research Institutes, National Taiwan University, and Baylor University. THRV is based (substantially) on the parent SAPPHIRe study, along with additional population-based and hospital-based cohorts. Whole genome sequencing (WGS) for the Trans-Omics in Precision Medicine (TOPMed) program was supported by the National Heart, Lung and Blood Institute (NHLBI). The Rare Variants for Hypertension in Taiwan Chinese (THRV) is supported by the National Heart, Lung, and Blood Institute (NHLBI) grant (R01HL111249) and its participation in TOPMed is supported by an NHLBI supplement (R01HL111249-04S1). SAPPHIRe was supported by NHLBI grants (U01HL54527, U01HL54498) and Taiwan funds, and the other cohorts were supported by Taiwan funds.

#### Women’s Health Initiative (WHI, n=8,263):

*TOPMed dbGaP accession#: phs001237, Parent dbGaP accession#: phs000200, Sequencing Center: Broad Institute of MIT and Harvard.*

The Women's Health Initiative (WHI) is a large study of postmenopausal women’s health investigating risk factors for cancer, CVD, age-related fractures and chronic disease. It began in 1993 as a set of randomized controlled clinical trials (CT) and an observational study (OS). Specifically, the CT (n=68,132) included three overlapping components: The Hormone Therapy (HT) Trials (n=27,347), Dietary Modification (DM) Trial (n=48,835), and Calcium and Vitamin D (CaD) Trial (n=36,282). Eligible women could be randomized into as many as all three CTs components. Women who were ineligible or unwilling to join the CT were then invited to join the OS (n=93,676)^33,34^. WHI is a case control study with ~5000 strokes and >2000 CHD samples. Whole genome sequencing (WGS) for the Trans-Omics in Precision Medicine (TOPMed) program was supported by the National Heart, Lung and Blood Institute (NHLBI). The WHI program is funded by the National Heart, Lung, and Blood Institute, National Institutes of Health, U.S. Department of Health and Human Services through contracts HHSN268201600018C, HHSN268201600001C, HHSN268201600002C, HHSN268201600003C, and HHSN268201600004C.

### TOPMed Omics Support table

| **TOPMed Accession #** | **TOPMed Project** | **Parent Study** | **TOPMed Phase** | **Omics Center** | **Omics Support** | **Omics Type** |
| --- | --- | --- | --- | --- | --- | --- |
| **phs000956** | Amish | Amish | 1 | Broad Genomics | 3R01HL121007-01S1 | WGS |
| **phs001211** | AFGen | ARIC AFGen | 1 | Broad Genomics | 3R01HL092577-06S1 | WGS |
| **phs001211** | VTE | ARIC | 2 | Baylor | 3U54HG003273-12S2 / HHSN268201500015C | WGS |
| **phs001644** | AFGen | BioMe AFGen | 2.4 | MGI | 3UM1HG008853-01S2 | WGS |
| **phs001644** | BioMe | BioMe | 3 | MGI | HHSN268201600037I | WGS |
| **phs001644** | BioMe | BioMe | 3 | Baylor | HHSN268201600033I | WGS |
| **phs001612** | CARDIA | CARDIA | 3 | Baylor | HHSN268201600033I | WGS |
| **phs000954** | CFS | CFS | 3.5 | NWGC | HHSN268201600032I | WGS |
| **phs000954** | CFS | CFS | 1 | NWGC | 3R01HL098433-05S1 | WGS |
| **phs001368** | CHS | CHS | 3 | Baylor | HHSN268201600033I | WGS |
| **phs001368** | CHS | CHS | 5.5-5.6; 6 backfill | Broad Genomics | HHSN268201600034I | WGS |
| **phs001368** | VTE | CHS VTE | 2 | Baylor | 3U54HG003273-12S2 / HHSN268201500015C | WGS |
| **phs001412** | AA_CAC | DHS | 2 | Broad Genomics | HHSN268201500014C | WGS |
| **phs000974** | AFGen | FHS AFGen | 1 | Broad Genomics | 3R01HL092577-06S1 | WGS |
| **phs000974** | FHS | FHS | pilot; 4.5; 5.5-5.6; 6 backfill | Broad Genomics | HHSN268201600034I | WGS |
| **phs000974** | FHS | FHS | 1 | Broad Genomics | 3U54HG003067-12S2 | WGS |
| **phs001218** | AA_CAC | GeneSTAR AA_CAC | 2 | Broad Genomics | HHSN268201500014C | WGS |
| **phs001218** | GeneSTAR | GeneSTAR | 2 | Psomagen | 3R01HL112064-04S1 | WGS |
| **phs001218** | GeneSTAR | GeneSTAR | legacy | Illumina | R01HL112064 | WGS |
| **phs001345** | HyperGEN_GENOA | GENOA | 2 | NWGC | 3R01HL055673-18S1 | WGS |
| **phs001345** | AA_CAC | GENOA AA_CAC | 2 | Broad Genomics | HHSN268201500014C | WGS |
| **phs001217** | GenSalt | GenSalt | 2 | Baylor | HHSN268201500015C | WGS |
| **phs001359** | GOLDN | GOLDN | 2 | NWGC | 3R01HL104135-04S1 | WGS |
| **phs001395** | HCHS_SOL | HCHS_SOL | 3 | Baylor | HHSN268201600033I | WGS |
| **phs001293** | HyperGEN_GENOA | HyperGEN | 2 | NWGC | 3R01HL055673-18S1 | WGS |
| **phs000964** | JHS | JHS | 1 | NWGC | HHSN268201100037C | WGS |
| **phs001416** | AA_CAC | MESA AA_CAC | 2 | Broad Genomics | HHSN268201500014C | WGS |
| **phs001416** | MESA | MESA | 2 | Broad Genomics | 3U54HG003067-13S1 | WGS |
| **phs001062** | AFGen | MGH_AF | 1 | Broad Genomics | 3R01HL092577-06S1 | WGS |
| **phs001062** | AFGen | MGH_AF | 1.4; 1.5; 2.4 | Broad Genomics | 3U54HG003067-12S2 / 3U54HG003067-13S1; 3U54HG003067-12S2 / 3U54HG003067-13S1; 3UM1HG008895-01S2 | WGS |
| **phs001215** | SAFS | SAFS | legacy | Illumina | R01HL113322 | WGS |
| **phs001215** | SAFS | SAFS | 1 | Illumina | 3R01HL113323-03S1 | WGS |
| **phs000972** | Samoan | Samoan | 2 | NYGC Genomics | HHSN268201500016C | WGS |
| **phs000972** | Samoan | Samoan | 1 | NWGC | HHSN268201100037C | WGS |
| **phs001387** | THRV | THRV | 2 | Baylor | 3R01HL111249-04S1 / HHSN26820150015C | WGS |
| **phs001237** | WHI | WHI | 2 | Broad Genomics | HHSN268201500014C | WGS |

Baylor = Baylor College of Medicine Human Genome Sequencing Center

Broad Genomics = Broad Institute Genomics Platform

Broad Metabolomics = Broad Institute and Beth Israel Metabolomics Platform

Illumina = Illumina

Keck MGC = Keck Molecular Genomics Core Facility

MGI = McDonnell Genome Institute

NWGC = Northwest Genomics Center

NYGC Genomics = New York Genome Center Genomics

Psomagen = Psomagen

### Supplementary Text: TOPMed lipids working group

Gonçalo Abecasis, Donna K. Arnett, Stella Aslibekyan, Tim Assimes, Elizabeth Atkinson, Christie Ballantyne, Wei Bao, Amber Beitelshees, Romit Bhattacharya, Larry Bielak, Joshua Bis, Corneliu Bodea, Eric Boerwinkle, Donald W. Bowden, Jennifer Brody, Brian Cade, Sarah Calvo, Jenna Carlson, I-Shou Chang, Yii-Der Ida Chen, So Mi Cho, Seung Hoan Choi, Ren-Hua Chung, Adolfo Correa, L. Adrienne Cupples, Coleen Damcott, Paul de Vries, Ana F. Diallo, Ron Do, Jacqueline Dron, Amanda Elliott, Hilary Finucane, Caitlin Floyd, Mao Fu, Andrea Ganna, Dawei Gong, Sarah Graham, Mary Haas, Bernhard Haring, Jiang He, Scott Heemann, Blanca Himes, James Hixson, Marguerite Ryan Irvin, Gail Jarvik, Jicai Jiang, Roby Joehanes, Paule Valery Joseph, Goo Jun, Rita Kalyani, Masahiro Kanai, Sharon Kardia, Sekar Kathiresan, Amit Khera, Sumeet Khetarpal, Derek Klarin, Charles Kooperberg, Satoshi Koyama, Brian Kral, Leslie Lange, Cathy Laurie, Rozenn Lemaitre, Zilin Li, Xihao Li, Changwei Li, Xihong Lin, Yingchang Lu, Michael Mahaney, Ani Manichaikul, Lisa Martin, Rasika Mathias, Ravi Mathur, Stephen McGarvey, John McLenithan, Julie Mikulla, Amy Miller, Braxton D. Mitchell, May E. Montasser, Vamsi Mootha, Andrew Moran, Alanna C. Morrison, Tetsushi Nakao, Pradeep Natarajan, Kari North, Jeff O’Connell, Christopher O’Donnell, Nicholette Palmer, Kaavya Paruchuri, Aniruddh Patel, Gina Peloso, James Perry, Ulrike Peters, Mary Pettinger, Patricia Peyser, James Pirruccello, Toni Pollin, Michael Preuss, Bruce Psaty, Susan Redline, Robert Reed, Alex Reiner, Stephen Rich, Samantha Rosenthal, Jerome Rotter, Margaret Sunitha Selvaraj, Wayne Hui-Heng Sheu, Jennifer Smith, Tamar Sofer, Adrienne M. Stilp, Shamil R. Sunyaev, Ida Surakka, Carole Sztalryd, Hua Tang, Kent D. Taylor, Mark Trinder, Michael Tsai, Md Mesbah Uddin, Sarah Urbut, Eric Van Buren, Marie Verbanck, Ann Von Holle, Heming Wang, Yuxuan Wang, Kerri Wiggins, John Wilkins, Cristen Willer, James Wilson, Brooke Wolford, Huichun Xu, Lisa Yanek, Zhi Yu, Norann Zaghloul, Seyedeh Maryam Zekavat, Jingwen Zhang, Ying Zhou

### Supplementary Text: TOPMed Study specific grant acknowledgements

The Amish studies were supported by NIH grants R01 AG18728, U01 HL072515, R01 HL088119, R01 HL121007, and P30 DK072488. The Atherosclerosis Risk in Communities (ARIC) study has been funded in whole or in part with Federal funds from the National Heart, Lung, and Blood Institute, National Institutes of Health, Department of Health and Human Services (contract numbers HHSN268201700001I, HHSN268201700002I, HHSN268201700003I, HHSN268201700004I and HHSN268201700005I). The authors thank the staff and participants of the ARIC study for their important contributions. The Mount Sinai BioMe Biobank (BioMe) has been supported by The Andrea and Charles Bronfman Philanthropies and in part by Federal funds from the NHLBI and NHGRI (U01HG00638001; U01HG007417; X01HL134588). Coronary Artery Risk Development in Young Adults (CARDIA) Study (phs001612) was performed at the Baylor College of Medicine Human genome Sequencing Center (contract HHSN268201600033I). Core support including centralized genomic read mapping and genotype calling, along with variant quality metrics and filtering were provided by the TOPMed Informatics Research Center (3R01HL-117626-02S1; contract HHSN268201800002I). Core support including phenotype harmonization, data management, sample-identity QC, and general program coordination were provided by the TOPMed Data Coordinating Center (R01HL-120393; U01HL-120393; contract HHSN268201800001I). We gratefully acknowledge the studies and participants who provided biological samples and data for TOPMed. The Coronary Artery Risk Development in Young Adults Study (CARDIA) is conducted and supported by the National Heart, Lung, and Blood Institute (NHLBI) in collaboration with the University of Alabama at Birmingham (HHSN268201800005I & HHSN268201800007I), Northwestern University (HHSN268201800003I), University of Minnesota (HHSN268201800006I), and Kaiser Foundation Research Institute (HHSN268201800004I). Cleveland Family Study (CFS) is supported by grants from the NHLBI (HL046389, HL113338, and 1R35HL135818). Cardiovascular Health Study (CHS) was supported by contracts HHSN268201200036C, HHSN268200800007C, HHSN268201800001C, N01HC55222, N01HC85079, N01HC85080, N01HC85081, N01HC85082, N01HC85083, N01HC85086, 75N92021D00006, and grants U01HL080295 and U01HL130114 from the National Heart, Lung, and Blood Institute (NHLBI), with additional contribution from the National Institute of Neurological Disorders and Stroke (NINDS). Additional support was provided by R01AG023629 from the National Institute on Aging (NIA). A full list of principal CHS investigators and institutions can be found at CHS-NHLBI.org. The content is solely the responsibility of the authors and does not necessarily represent the official views of the National Institutes of Health. Diabetes Heart Study (DHS) was supported by HL92301, HL67348, NS058700, AR48797, DK071891, AG058921, the General Clinical Research Center of the Wake Forest University School of Medicine (RR07122, HL085989), the American Diabetes Association, and a pilot grant from the Claude Pepper Older Americans Independence Center of Wake Forest University Health Sciences (AG10484). Framingham Heart Study (FHS) acknowledges the support of contracts NO1-HC-25195 and HHSN268201500001I from the National Heart, Lung and Blood Institute and grant supplement R01 HL092577-06S1 for this research. WGS for “NHLBI TOPMed: Whole Genome Sequencing and Related Phenotypes in the Framingham Heart Study” (phs000974) was performed at the Broad Institute of MIT and Harvard (HHSN268201500014C, 3R01HL092577-06S1, and 3U54HG003067-12S2). We also acknowledge the dedication of the FHS study participants without whom this research would not be possible. Genetic Studies of Atherosclerosis Risk (GeneSTAR) was supported by grants from the National Institutes of Health/National Heart, Lung, and Blood Institute (U01 HL72518, HL087698, HL49762, HL58625, HL071025, HL112064), the National Institutes of Health/National Institute of Nursing Research (NR0224103), and by a grant from the National Institutes of Health/National Center for Research Resources (M01-RR000052) to the Johns Hopkins General Clinical Research Center. Genetic Epidemiology Network of Arteriopathy (GENOA) was supported by the National Heart, Lung and Blood Institute (HL054457, HL054464, HL054481, HL087660, and HL119443) of the National Institutes of Health. Genetic Epidemiology Network of Salt Sensitivity (GenSalt) was supported by research grants (U01HL072507, R01HL087263, and R01HL090682) from the National Heart, Lung and Blood Institute, National Institutes of Health, Bethesda, MD. Genetics of Lipid-Lowering Drugs and Diet Network (GOLDN) biospecimens, baseline phenotype data, and intervention phenotype data were collected with funding from National Heart, Lung and Blood Institute (NHLBI) grant U01 HL072524. The Hispanic Community Health Study/Study of Latinos (HCHS- SOL) was carried out as a collaborative study supported by contracts from the National Heart, Lung, and Blood Institute (NHLBI) to the University of North Carolina (N01- HC65233), University of Miami (N01-HC65234), Albert Einstein College of Medicine (N01-HC65235), Northwestern University (N01-HC65236), and San Diego State University (N01-HC65237). The Hypertension Genetic Epidemiology Network and Genetic Epidemiology Network of Arteriopathy (HyperGEN) Study is part of the National Heart, Lung, and Blood Institute (NHLBI) Family Blood Pressure Program; collection of the data represented here was supported by grants U01 HL054472 (MN Lab), U01 HL054473 (DCC), U01 HL054495 (AL FC), and U01 HL054509 (NC FC). The HyperGEN: Genetics of Left Ventricular Hypertrophy Study was supported by NHLBI grant R01 HL055673 with whole-genome sequencing made possible by supplement - 18S1. The Jackson Heart Study (JHS) is supported and conducted in collaboration with Jackson State University (HHSN268201800013I), Tougaloo College (HHSN268201800014I), the Mississippi State Department of Health (HHSN268201800015I) and the University of Mississippi Medical Center (HHSN268201800010I, HHSN268201800011I and HHSN268201800012I) contracts from the National Heart, Lung, and Blood Institute (NHLBI) and the National Institute on Minority Health and Health Disparities (NIMHD). The authors also wish to thank the staffs and participants of the JHS. Multi-Ethnic Study of Atherosclerosis (MESA) and the MESA SHARe projects are conducted and supported by the National Heart, Lung, and Blood Institute (NHLBI) in collaboration with MESA investigators. Support for MESA is provided by contracts 75N92020D00001, HHSN268201500003I, N01-HC-95159, 75N92020D00005, N01-HC-95160, 75N92020D00002, N01-HC-95161, 75N92020D00003, N01-HC-95162, 75N92020D00006, N01-HC-95163, 75N92020D00004, N01-HC-95164, 75N92020D00007, N01-HC-95165, N01-HC-95166, N01-HC-95167, N01-HC-95168, N01-HC-95169, UL1-TR-000040, UL1-TR-001079, and UL1-TR-001420. Funding for SHARe genotyping was provided by NHLBI Contract N02-HL-64278. Genotyping was performed at Affymetrix (Santa Clara, California, USA) and the Broad Institute of Harvard and MIT (Boston, Massachusetts, USA) using the Affymetrix Genome-Wide Human SNP Array 6.0. Also supported in part by NHLBI CHARGE Consortium Contract HL105756. The provision of genotyping data was supported in part by the National Center for Advancing Translational Sciences, CTSI grant UL1TR001881, and the National Institute of Diabetes and Digestive and Kidney Disease Diabetes Research Center (DRC) grant DK063491 to the Southern California Diabetes Endocrinology Research Center. Infrastructure for the CHARGE Consortium is supported in part by the National Heart, Lung, and Blood Institute (NHLBI) grant R01HL105756. The Massachusetts General Hospital Atrial Fibrillation Study (MGH-AF) was supported by grants to Dr. Ellinor from the Fondation Leducq (14CVD01), the National Institutes of Health to Dr. Ellinor (1RO1HL092577, R01HL128914, K24HL105780) and Dr. Lubitz (1R01HL139731) and by grants from the American Heart Association to Dr. Ellinor (18SFRN34110082) and to Dr. Lubitz (18SFRN34250007). San Antonio Family Study (SAFS) was supported in part by National Institutes of Health (NIH) grants R01 HL045522, MH078143, MH078111 and MH083824; and whole genome sequencing of SAFS subjects was supported by U01 DK085524 and R01 HL113323. We are very grateful to the participants of the San Antonio Family Study for their continued involvement in our research programs. Samoan Adiposity Study (SAS) was funded by NIH grant R01-HL093093. We thank the Samoan participants of the study and local village authorities. We acknowledge the support of the Samoan Ministry of Health and the Samoa Bureau of Statistics for their support of this research. The Rare Variants for Hypertension in Taiwan Chinese (THRV) is supported by the National Heart, Lung, and Blood Institute (NHLBI) grant (R01HL111249) and its participation in TOPMed is supported by an NHLBI supplement (R01HL111249-04S1). SAPPHIRe was supported by NHLBI grants (U01HL54527, U01HL54498) and Taiwan funds, and the other cohorts were supported by Taiwan funds. The Women’s Health Initiative (WHI) program is funded by the National Heart, Lung, and Blood Institute, National Institutes of Health, U.S. Department of Health and Human Services through contracts HHSN268201600018C, HHSN268201600001C, HHSN268201600002C, HHSN268201600003C, and HHSN268201600004C. The Centers for Common Disease Genomics (CCDG) program was supported by NHGRI and NHLBI, and whole genome sequencing was performed at the Baylor College of Medicine Human Genome Sequencing Center (UM1 HG008898 and R01HL059367).

### Supplementary Text: FHS RNA-seq Protocol

#### Study participants

This study included 1505 participants from the FHS Third Generation cohorts^35^. Blood samples for RNA seq were collected from Third Generation participants who attended the second examination cycle (2008–2011). Protocols for participant examinations and collection of genetic materials were approved by the Institutional Review Board at Boston Medical Center. All participants provided written, informed consent for genetic studies. All research was performed in accordance with relevant guidelines/regulations.

#### RNA-seq data collection, quality control, and data adjustment

The process of collection and isolation of RNA from whole blood was described previously^36^. All RNA samples were sequenced by an NHLBI TOPMed program^37^ reference laboratory (Northwest Genomics Center) following the TOPMed RNA-seq protocol. All RNS-seq data were processed by the University of Washington. The raw reads (in FASTQ files) were aligned using the GRCh38 reference build to generate BAM files. RNA-SeQC^38^ was used for processing of RNA-seq data by the TOPMed RNA-seq pipeline to derive standard quality control metrics from aligned reads. Gene-level expression quantification was provided as read counts and transcripts per million (TPM). GENCODE v30 annotation was used for annotating gene-level expression.

We performed the trimmed mean of M values (TMM) normalization on the gene read counts of RNA-seq data using the *edgeR* R/Bioconductor package^39,40^. We removed the lowly expressed transcripts that have a SD equal to 0. To minimize confounding, expression residuals were generated by regressing log2(TMM+1) values on technical covariates including year of blood collection, batch (sequencing machine and time, plate and well), and RNA concentration.

#### Predicted complete blood count (CBC)

Because 80% of the participants in this study had directly measured cell count variables and only 20% received imputed variables, partial least squares (PLS) method^41^ was used to create predicted complete blood count data based on the RNA-seq data. To improve the prediction, we set the Basophil percentage (BA_PER) that is greater than 3 as missing. We performed a partial least squares (PLS) prediction method with three-fold cross-validation (2/3 samples for training and 1/3 for validation) to impute these blood cell components using gene expression from RNA-seq^42^. We then tested the accuracy in the testing dataset. Prediction accuracy (R-squared) varied across blood component: WBC: 58%, platelet: 27%, neutrophil percentage: 82%, lymphocyte percentage: 85%, monocyte percentage: 77%, eosinophil percentage: 87%, basophil percentage: 32%.

#### Statistical analysis

We fitted a linear mixed effects model for the residuals of the TMM normalized log2 transformed counts data and the lipid phenotypes adjusting for predicted complete blood count (CBC), constructed surrogate variables (SVs), sex, age, and family structure as variance-covariance matrix using *GENESIS* R/Bioconductor package^43^. Surrogate variables (SVs) are covariates constructed directly from gene expression data to adjust for unknown, unmodeled, or latent sources of noise^44^. We estimated the SVs from expression residuals and each lipid phenotype using the R/Bioconductor *sva* package^45^. For each association, we collected the effect estimate (*β*), T-statistics, and *P* values.

### Supplementary Figures: Legends

#### Supplementary Figure 1. Sensitivity analysis comparing using all rare lncRNA variants versus using only rare exonic or splicing lncRNA variants to define the test units.

Each dot represents the -log_10_(STAAR-O *P* value). The red dashed line is the diagonal line. **a**, Rare lncRNA variant sets for LDL-C. **b**, Rare lncRNA variant sets for TC. **c**, Rare lncRNA variant sets for HDL-C. **d**, Rare lncRNA variant sets for TG. STAAR – variant-Set Test for Association using Annotation information; HDL-C – High-Density Lipoprotein Cholesterol; LDL-C – Low-Density Lipoprotein Cholesterol; TC – Total Cholesterol; TG – Triglycerides.

#### Supplementary Figure 2. Results for the 61 lncRNA-lipid associations that remained significant (STAAR-O P value < 6.0e-04) in the conditional analysis adjusting for known lipid-associated GWAS variants.

The lncRNA genes are ordered based on chromosomes, followed by their genomic positions. Dots in red, blue, green, and purple represent the -log10(STAAR-O *P* value) of the STAAR unconditional analysis, STAAR conditional analysis adjusting on known lipid-associated GWAS variants, STAAR conditional analysis adjusting on closest gene and nearby lipid protein coding genes, and replication analysis adjusting on known lipid GWAS variants in UKBB, respectively. The black dashed line is the Bonferroni correction level 0.05/61 = 8.2e-04. **a**, Rare lncRNA variant sets for LDL-C. **b**, Rare lncRNA variant sets for TC. **c**, Rare lncRNA variant sets for HDL-C. **d**, Rare lncRNA variant sets for TG. STAAR – variant-Set Test for Association using Annotation information; HDL-C – High-Density Lipoprotein Cholesterol; LDL-C – Low-Density Lipoprotein Cholesterol; TC – Total Cholesterol; TG – Triglycerides. UKBB – UK BioBank.

#### Supplementary Figure 3. Comparison of results for the unconditional analysis and all the conditional analyses.

Each dot is the -log_10_(STAAR-O *P* value). The red dashed line is the diagonal line. **a**, Unconditional vs Adjusting for known lipid GWAS variants. **b**, Unconditional vs Adjusting for rare nonsynonymous variants within nearby protein coding genes. **c**, Unconditional vs Adjusting for rare synonymous variants within nearby protein coding genes. **d**, Unconditional vs Adjusting for rare pLoF variants within nearby protein coding genes. STAAR – variant-Set Test for Association using Annotation information. P (STAAR-O *P* value of unconditional analysis), P_GWAS_cond (STAAR-O *P* value of conditional analysis adjusting for known lipid-associated GWAS variants), P_non_synonymous_cond (STAAR-O *P* value of conditional analysis adjusting for rare nonsynonymous variants within the closest gene and nearby lipid monogenic genes), P_synonymous_cond (STAAR-O *P* value of conditional analysis adjusting for rare synonymous variants within the closest gene and nearby lipid monogenic genes), P_plof_cond (STAAR-O *P* value of conditional analysis adjusting for rare pLoF variants within the closest gene and nearby lipid monogenic genes)

### Supplementary Tables: Legends

#### Supplementary Table 1: Study-specific sample sizes and baseline characteristics of 66, 329 participants in TOPMed freeze 8 data from 21 cohorts.

Samples are tabulated based on gender, ancestral groups, sequencing centers, and lipid treatment (percentage of samples) for each cohort. Distribution of mean [SD] for sex, unadjusted non-transformed lipid concentration of HDL-C, LDL-C, TC and TG in full sample group and stratified groups based on gender are provided. TG concentration is summarized as median [IQR] for all samples separately. Lipid concentrations are in units of mg/dl. Cohorts with no values for any specific columns are represented as NA. Sequencing centers: Baylor: Baylor College of Medicine Human Genome Sequencing Center, Broad: Broad Institute of MIT and Harvard, Illumina: Illumina Genomic Services, Macrogen: PSOMAGEN (formerly Macrogen), NYGC: New York Genome Center, UW: McDonnell Genome Institute (MGI) at Washington University, WASHU: Northwest Genomics Center. HDL-C – High-Density Lipoprotein Cholesterol; LDL-C – Low-Density Lipoprotein Cholesterol; TC – Total Cholesterol; TG – Triglycerides

#### Supplementary Table 2: STAAR lncRNA analysis results for HDL-C, LDL-C, TC and TG using the TOPMed Freeze 8 WGS data.

Results for the 83 significant lncRNA genes (unconditional STAAR-O *P* value < 4.5e-07) are presented in the table. The unconditional significant threshold for lncRNA genes was defined by the multiple comparisons using the Bonferroni correction and the effective number of tests, i.e., 0.05/111,550 = 4.5e-07. CHR(chromosome), HDL-C (High-Density Lipoprotein Cholesterol), LDL-C (Low-Density Lipoprotein Cholesterol), TC (Total Cholesterol), TG (Triglycerides), GWAS SNV Adjusted (adjusted common and low frequency variants in conditional analysis); Protein Coding Genes Adjusted (adjusted rare nonsynonymous variants within the closest gene and nearby lipid monogenic genes), #SNV (number of rare variants, MAF < 1%), #SNV.UKBB (number of rare variants in replication cohort, MAF < 1%), P (STAAR-O *P* value of unconditional analysis), P_GWAS_cond (STAAR-O *P* value of conditional analysis adjusting for known lipid-associated GWAS variants), P_non_synonymous_cond (STAAR-O *P* value of conditional analysis adjusting for rare nonsynonymous variants within the closest gene and nearby lipid monogenic genes), P_synonymous_cond (STAAR-O *P* value of conditional analysis adjusting for rare synonymous variants within the closest gene and nearby lipid monogenic genes), P_plof_cond (STAAR-O *P* value of conditional analysis adjusting for rare pLoF variants within the closest gene and nearby lipid monogenic genes), P_gwas_cond_UKBB (STAAR-O *P* value of replication analysis adjusting for known lipid-associated GWAS variants in UK Biobank data).

#### Supplementary Table 3: Baseline characteristics of replication cohort.

Sample sizes, gender distributions, ancestry distributions, and mean ages of individuals in the replication cohort are provided. Mean (standard deviation) of unadjusted and non-transformed lipid concentrations are presented.

#### Supplementary Table 4: Gene expression and lipid traits associations of the significant associated GENCODE lncRNA genes identified from the TOPMed WGS study.

The significant threshold was defined by the multiple comparisons using the Bonferroni correction, that is, 0.05/12=4.17E-03. LDL-C (low-density lipoprotein cholesterol); HDL-C (high-density lipoprotein cholesterol); TG (triglycerides); TC (total cholesterol).

#### Supplementary Table 5: Individual and integrative variant functional annotations used in the STAAR framework.

Tissue-specific annotations (liver) DNase, H3K4me3, H3K27ac and H3K27me3 are from ENCODE (https://www.encodeproject.org/report/?type=Experiment). Each annotation Principal Component (aPC) is the first PC calculated from the set of individual functional annotations that measure similar biological function. These aPCs are then transformed into the PHRED-scaled scores (-10*log10(rank/total)) for each variant across the genome. For aPC-LocalDiversity, we consider two separate scores generated by ranks from both directions.

#### Supplementary Table 6: Global Lipids Genetics Consortium multi-ancestry meta-analysis index variants that are associated with one or more lipid levels.

The positions of SNV were lifted over to genome build 38.

### Supplementary Figures

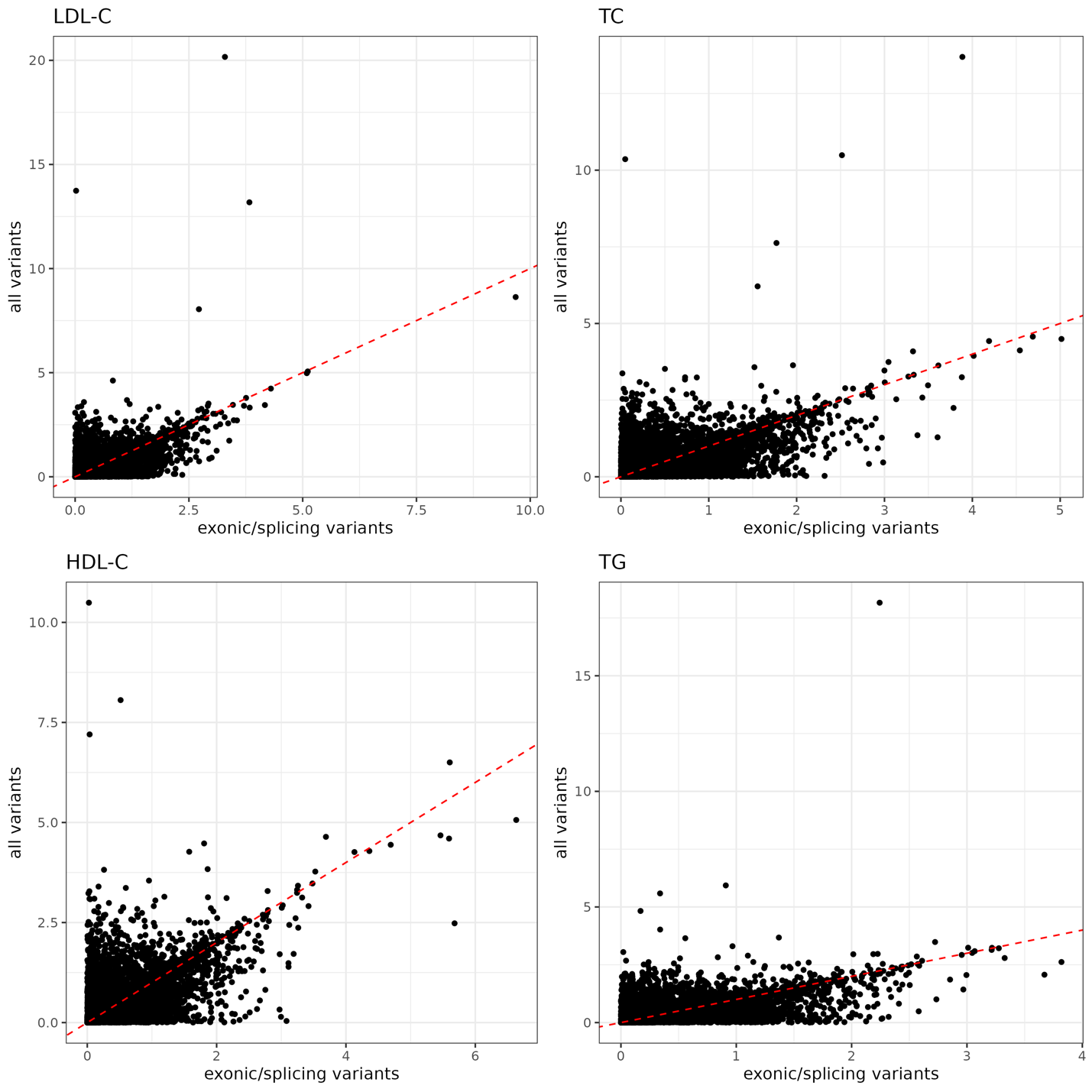

#### Supplementary Figure 1. Sensitivity analysis comparing using all rare lncRNA variants versus using only rare exonic or splicing lncRNA variants to define the test units.

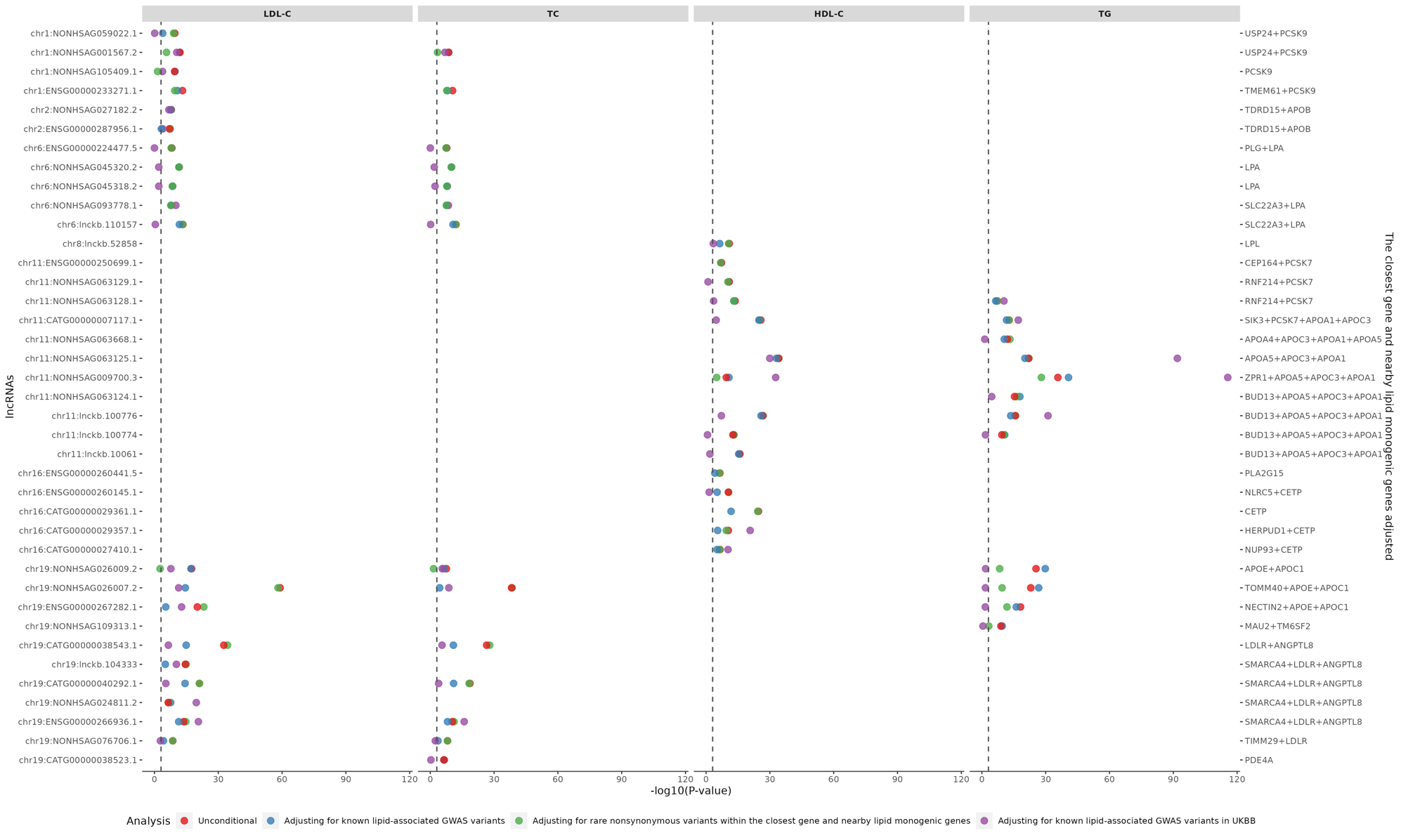

#### Supplementary Figure 2. Results for the 61 lncRNA-lipid associations that remained significant (STAAR-O P value < 6.0e-04) in the conditional analysis adjusting for known lipid-associated GWAS variants.

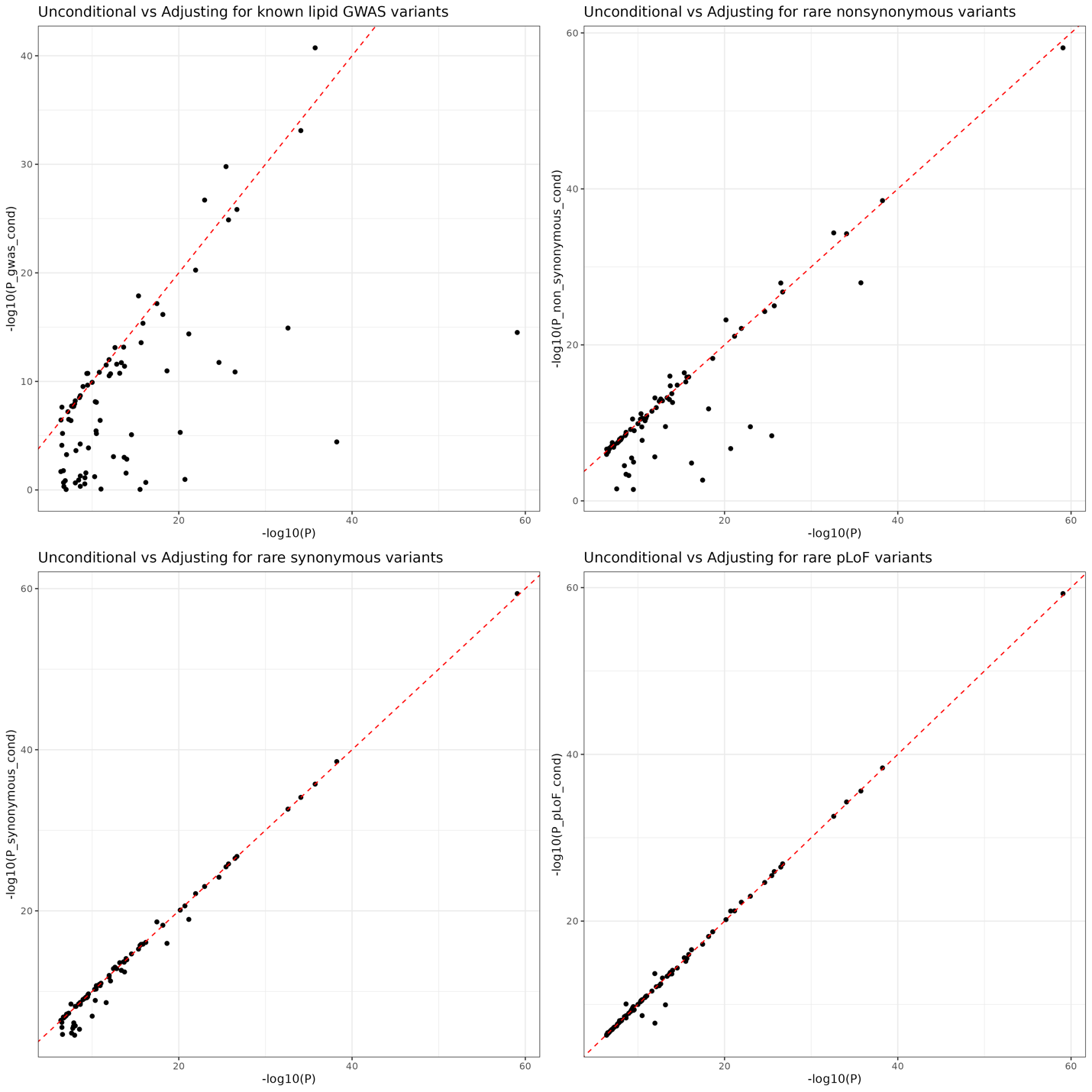

#### Supplementary Figure 3. Comparison of results for the unconditional analysis versus all the conditional analyses.
